# Motor Cortex Coverage Predicts Signal Strength of a Stentrode Endovascular Brain-Computer Interface

**DOI:** 10.1101/2025.09.19.25335875

**Authors:** Hunter R. Schone, Peter Yoo, Adam Fry, Nikole Chetty, Abbey Sawyer, Cara Herbers, Fang Liu, Chan Hong Moon, Katya Hill, Shahram Majidi, Noam Y. Harel, Raul G. Nogueira, Elad Levy, David F. Putrino, David Lacomis, Thomas J. Oxley, Douglas J. Weber, Jennifer L. Collinger

## Abstract

Brain-computer interfaces (BCIs) are an emerging assistive technology for individuals with motor impairments, enabling the command of digital devices using neural signals. The Stentrode BCI is an implant, positioned within the brain’s neurovasculature, that can record movement-related electrocortical activity. Over 5 years, 10 participants (8 amyotrophic lateral sclerosis, 1 primary lateral sclerosis, 1 brainstem stroke) have been implanted with a Stentrode BCI and significant inter-participant variability has been observed in the recorded motor signal strength. This variability warrants a critical investigation to characterize potential predictors of signal strength to promote more successful BCI control in future participants. Therefore, we investigated the relationship between Stentrode BCI motor signal strength and a variety of user-specific factors: (1) clinical status, (2) pre-implant functional activity, (3) peri-implant neuroanatomy, (4) peri-implant neurovasculature, and (5) Stentrode device integrity. Data from 10 implanted participants, including clinical demographics, pre- and post-implant neuroimaging and longitudinal Stentrode BCI motor signal assessments were acquired over a year. Across all potential predictors, the strongest predictor of Stentrode motor signal strength was the degree to which the Stentrode BCI’s deployment position overlapped with primary motor cortex (M1). These findings highlight the importance of targeting M1 during device deployment and, more generally, provides a scientific framework for investigating the role of user-specific factors on BCI device outcomes.

## Introduction

Since the first chronic implant in 1998, brain-computer interfaces (BCI) have steadily emerged as an increasingly feasible class of assistive technology^1^. Typically designed for individuals with motor impairments (e.g., due to spinal cord injury, stroke, or amyotrophic lateral sclerosis), BCIs translate brain signals into digital commands, circumventing impaired peripheral motor pathways to facilitate control of digital and physical end-effectors (e.g., computer cursors, robotic limbs), enable digital communication, and offer social engagement via online platforms^2^. Existing BCI systems can record brain signals at different locations, with systems using electrodes placed on the surface of the scalp or electrodes that interface directly within brain tissue. Each interface site offers distinct trade-offs in signal fidelity, surgical invasiveness, long-term usability and widespread adoption^1^. Scalp electroencephalography BCIs take a noninvasive approach, but suffer from limited signal resolution and prolonged setup times^3^.

Subdural electrocorticography (ECoG) makes use of electrodes laid on the surface of the brain and intracortical microelectrode arrays are inserted directly into the cortical tissue. These techniques offer high-resolution neural recordings, but require a craniotomy, i.e., open-brain surgery, thus limiting widespread adoption. Additionally, electrodes implanted directly into brain tissue exhibit a decline in signal quality over time, due to a biological neuroinflammatory response in the brain^4^ and material degradation^5–8^. An emerging alternative interface, the Stentrode BCI (developed by *Synchron, USA*), is an endovascular stent-electrode array that is implanted into the brain’s vasculature using common endovascular procedures. To date, the Stentrode BCI has been placed in the superior sagittal sinus (SSS), a large vein which lies at the midline of the brain, between the hemispheres of the brain. Unlike subdural ECoG and intracortical devices, the deployment approach does not require a craniotomy. Further, by leveraging the location of the neurovasculature relative to the brain, the Stentrode BCI can record distant neural activity without penetrating the cortical surface and is therefore being investigated as an alternative approach to intracortical devices (Figure 1A).

**Figure 1.**
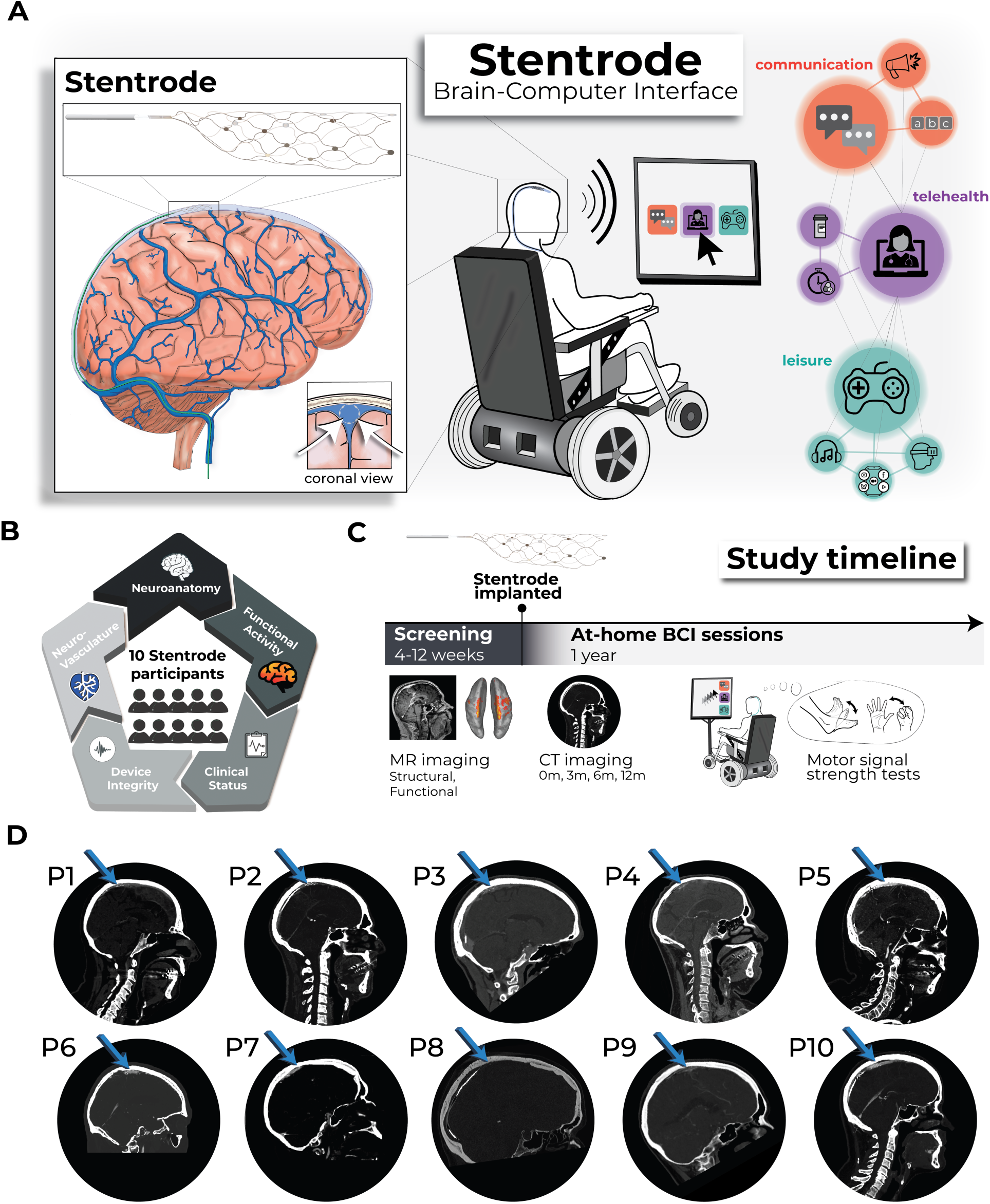
Study overview investigating participants implanted with a Stentrode BCI. **(A) Left –** llustration depicting the Stentrode BCI implanted within the superior saggital sinus.**Middle –** A Stentrode BCI user is depicted generating digital commands with their neural activity. **Right –** Real-world BCI applications include digital communication (orange), telehealth (purple) and leisure (teal). **(B)** Pooling data across 10 participants in the Stentrode BCI clinical trial, we tested which user-specific factors drive Stentrode BCI signal strength, including: neuroanatomy, pre-implant functional MRI activity, clinical condition, device integrity and neurovasculature. **(C)** Clinical trial timeline: pre-implant screening and at-home BCI sessions. **(D)** Post-implant CT scans of each participant at their 3-month timepoint illustrates the relative position of the Stentrode BCI location.

Despite the growing number of BCI technologies, only recently have at-home clinical trials begun to test BCI device usability, marking a critical shift in the focus from proof-of-concept demonstrations in controlled laboratory settings to real-world usability and performance evaluations (for a comprehensive review of all human BCI clinical trials^9^). Industry-led feasibility trials, such as those for the Stentrode BCI [Stentrode clinical trials (Clinicaltrials.gov): AUS (n=4): NCT03834857^10,11^; USA (n=6): NCT05035823], have investigated the safety of the implanted device and provided an opportunity to gather efficacy data, beginning to define for the field what BCI usability looks like in users’ daily lives. However, experience from at-home use of the Stentrode BCI—and similar observations from other BCI technologies—has highlighted a new challenge: BCI signal strength can vary substantially across participants^12–15^. Moving forward, it is necessary to identify which user-specific factors most reliably predict BCI signal strength for each device, whether related to a participant’s clinical status, device design, or where the BCI is implanted. This information is critical for designing pivotal clinical trials that drive the technology’s path to widespread clinical use. Further, considering the clinical risks for any BCI implantation procedure and the costs and resources required to implant a single BCI device, it is necessary to critically evaluate, at this stage, whether any user-specific factors can predict the inter-participant variability in Stentrode BCI motor signal strength. Crucially, this question–what factors best predict successful BCI use–extends beyond endovascular BCIs to the entire field, where predicting and optimizing BCI performance is going to be essential for the widespread clinical deployment of devices and selecting the best BCI technology for each patient’s neuroanatomy, clinical status and functional needs.

In the present work, we aimed to identify factors that may contribute to neural signal strength recorded in individuals who had been implanted with a Stentrode device. We hypothesize that several factors will contribute to the inter-participant variability in BCI motor signal strength, which we define as the amount of modulation in neural recordings during attempted movements^14^. Some of these user-specific factors are unique to endovascular BCIs and others are more general (Figure 1B). These include ***clinical status***, such as time since diagnosis and severity of motor impairment; ***pre-implant cortical function***, reflecting the extent to which users can activate motor cortex during attempted movements as measured by functional neuroimaging; ***peri-implant neuroanatomy***, encompassing the spatial proximity of the implant to cortical tissue and specific brain regions; ***peri-implant neurovasculature***, such as width of the SSS and its relative distance to the cortical surface; and ***device integrity***, specifically the number of active recording electrodes (Figure 1C). To address these open questions, across all participants historically implanted with a Stentrode BCI, we pooled datasets including clinical demographics, functional and structural neuroimaging (pre- and post-implant), and longitudinal Stentrode BCI motor signal strength recordings (n=10; Figure 1D). Combined, our investigation aimed to identify the strongest user-specific predictors of Stentrode BCI motor signal strength. Across all tested predictors, the most significant predictor of Stentrode BCI motor signal strength was how much the Stentrode BCI overlapped with motor cortex, demonstrating that successful endovascular BCI use critically depends on targeting primary motor cortex— highlighting the importance of precise neuroanatomical targeting for future clinical deployment.

## Results

Our pipeline involved first ***extracting the user-specific factors***, then ***quantifying Stentrode BCI motor signal strength*** during attempted movements across multiple sessions, and finally ***building predictive models*** to test which factors are associated with signal strength.

### Clinical status

Heterogeneity in disease state across study participants

Ten study participants with motor impairment were implanted with a Stentrode BCI. Nine participants were diagnosed with adult-onset motor neuron diseases [8 amyotrophic lateral sclerosis (ALS); 1 primary lateral sclerosis (PLS; P4); mean ± STD; 4.5 ± 2.8 years since diagnosis at the time of consent], and 1 participant (P8) was diagnosed with an arterial ischemic stroke in the brainstem (13.5 years since diagnosis at time of consent; see participant demographics in Supp. Table 1). Participants varied in their functional status at the time of implant (for qualitative descriptions of participant’s functional status, see Supp. Table 1). To quantify motor function, participants were graded on their ability to generate muscle contractions of different body-parts: the fingers, wrist, elbow, shoulder, hip, knee, ankle and toes. Participants varied in their manual muscle testing scores (^16,17^). We hypothesized that the progression of a participant’s motor neuron disease pathology or residual motor strength could be potential predictors of Stentrode BCI motor signal strength.

### Pre-implant functional neuroimaging

Preserved ability to activate primary motor cortex

Prior to implantation, all participants underwent functional and structural MRI scans (see Methods). The purpose of the functional MRI was to characterize each participant’s ability to functionally activate sensorimotor cortex during attempted movement. Participants needed to show some significant activation of motor cortex in order to be included in the study. During the functional MRI, participants were cued to perform single- or dual-ankle movements. The ankle was selected because ankle cortical activity is highly medial and superior^18^, making it the closest representation to the planned Stentrode BCI deployment site within the SSS. Projecting the ankle activation maps for each participant onto the cortical surface revealed widespread activation centered around sensorimotor cortices, though varying in breadth and strength across participants (Figure 2A). On average, across participants, the majority of the activity was within the boundaries of M1, somatosensory cortex (S1) and the supplementary motor area (SMA; Figure 2B-C). Further, there were stronger activations in M1 compared to S1 (paired Wilcoxon signed-rank test: W=47.0; *p*=0.04) and SMA (W=49.0; *p*=0.02). However, the spatial spread of significantly activated voxels (Z > 2.3) within each region was largely the same across regions (Supp. Figure 1A; M1 to S1: W=27.0; *p*=1.0; M1 to SMA: W=30; *p*=0.4; S1 to SMA: W=35.0; *p*=0.4).

**Figure 2.**
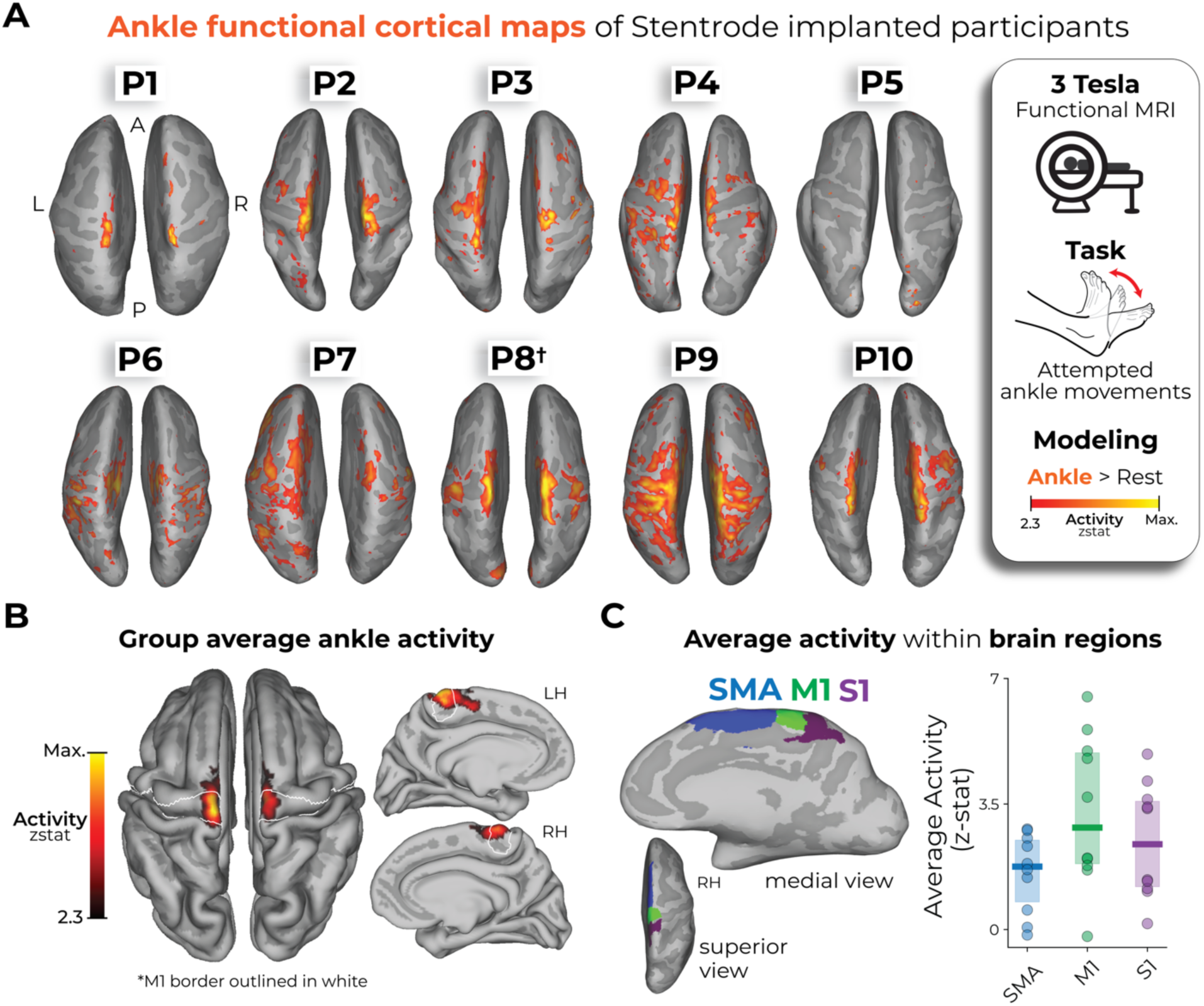
Pre-implant functional neuroimaging of Stentrode-implanted participants. **(A)** Ankle cortical maps of Stentrode-implanted participants. Using a 3 Tesla MRI scanner, participants were cued to perform ankle movements of either a single ankle or both ankles (see Methods). The functional activity for the ankle movement versus rest is projected onto each participant’s inflated cortical surface, depicting just the superior view. The activity is minimally thresholded at a z-statistic of 2.3. All participants were diagnosed with adult motor neuron diseases (ALS or PLS), except for P8 who was diagnosed with a pontine arterial ischemic stroke in the brainstem (depicted with a dagger symbol: †). **(B)** Averaging across participants, the group-level average ankle activation map is projected on a standard pial cortical surface. The primary motor cortex (M1; BA4) boundary is outlined in white. **(C)** Performing an ROI-specific analysis (averaged across both hemispheres), the M1, S1 and SMA ROIs were reduced in size to best capture the relevant ankle activation near the top of the cortical surface, closest to the Stentrode BCI deployment site. **Left** – An example visualization of a participant’s modified ROIs shown on an inflated, right hemisphere (RH) cortical surface. Across participants, M1 exhibited significantly stronger activity than SMA (paired Wilcoxon signed rank test: W=49.0, p_uncorr_=0.02) and S1 (W=47.0, p_uncorr_=0.04). **Right** – The average activity within each ROI for each participant is shown. Blue=SMA; green=M1; purple=S1.

We note P5 showed minimal functional activation compared to other participants, though there were still significantly activated voxels for ankle movements (see Supp. Figure 1A-B). We suspect that the reduced activation volume can be attributed to either poor task compliance during the scan (e.g., poor adherence to the attempted movement or unintended attempted movement during the rest period) and/or due to the late-stage progression of their ALS pathology (11 years since their diagnosis; see the *Discussion* section for more information). However, across all participants we saw no significant relationship between a participant’s ability to functionally activate motor cortex (average activation) and years since motor neuron disease diagnosis (*r_s_*=-0.50, *p*=0.91). Thus, despite motor neuron disease progression (or P8’s brainstem stroke), all participants were able to functionally activate motor cortex during attempted movement. While there is an obvious potential for selection bias given that the ability to activate motor cortex was part of the inclusion criteria, no participants who were screened were excluded for this reason in either clinical trial. However, some did repeat scans to test for significant activation. Aiming to leverage these preserved cortical motor representations to generate digital commands, all participants were then implanted with a Stentrode BCI (Figure 1D; see *Methods*).

### Structural neuroimaging

Variability in peri-implant neuroanatomy surrounding the Stentrode BCI

Next, we used the pre-implant structural MRI to generate high-resolution images of the cortical anatomy and neurovasculature architecture. After participants were implanted with the Stentrode BCI, CT scans were taken to visualize the location of the Stentrode BCI within the superior sagittal sinus (SSS). Each participant’s 3-month post-implant CT images (Figure 1D) were registered to their pre-implant structural T1w MRI. This allowed for visualization of each participant’s cortical surface reconstruction, neurovasculature and Stentrode BCI segmentations in a common coordinate space (Figure 3; see Supp. Figure 2 for an illustration of the method). Using this aligned data, our goal was to quantify how close the Stentrode BCI was to movement-related neural activity and anatomically-defined cortical regions (i.e., M1, S1, and SMA). We also quantified M1 cortical atrophy for the motor neuron disease participants, since motor neuron disease results in the death of motor neurons as the disease progresses^19,20^.

**Figure 3.**
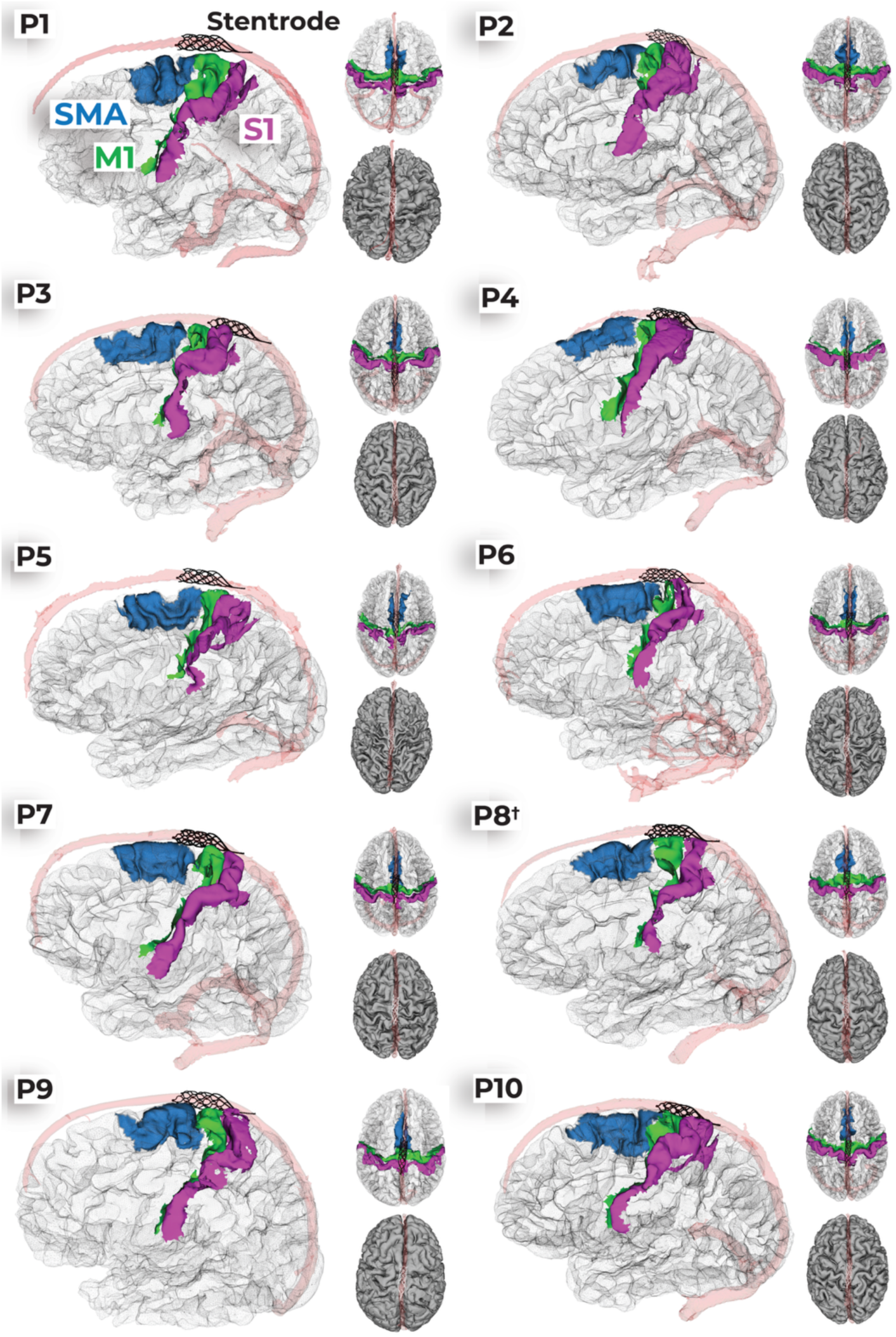
Stentrode BCI position for all participants. For each participant, there are 3 distinct visualizations of the implant position. First, there is a visualization of the sagittal (left) and axial (right top) views of an opaque wire-frame mesh of the cortical surfaces with vertices for specific brain regions colored (supplementary motor area=blue; primary motor cortex=green; somatosensory cortex=pink/purple) and the Stentrode model (black). Second, there is an axial view (right bottom) visualization of each participant’s pial cortical surfaces (grey) depicted with both the superior sagittal sinus segmentation (transparent red) and the Stentrode model (white). P8’s subject label includes a dagger symbol (†) to reflect their brainstem stroke.

First, we measured the distance between the Stentrode BCI and the cortical surface. Since individual electrodes are embedded at different points around the circumference of the stent scaffold—some closer to the cortex and others within the opposite wall of the SSS—we created a simplified approximation of its location. Specifically, we generated a 25mm-long line from the most rostral point of the Stentrode CT segmentation, extending caudally, centered within the SSS, and shaped to match its inferior curvature (Figure 4A-B). This approximation was used for all subsequent analyses. To account for cortical folding beneath the vasculature, we calculated (1) the average Stentrode-to-cortex distance across hemispheres and (2) the absolute minimum distance. Across participants, the Stentrode was, on average, 6.1 ± 1.5mm from the cortical surface, with the minimum distance of 3.7mm ± 1.2 (averaged across participants; Figure 4C). Though, because the array was modelled in the middle of the SSS, the reported values describe the mean electrode-to-cortex separation across the entire Stentrode, rather than the smallest possible clearance of its nearest contact. In addition to measuring distances to cortex, we quantified the width of the SSS vasculature segmentation around the Stentrode implant site, which averaged 7.1 ± 0.9 mm across participants (Supp. Figure 3).

**Figure 4.**
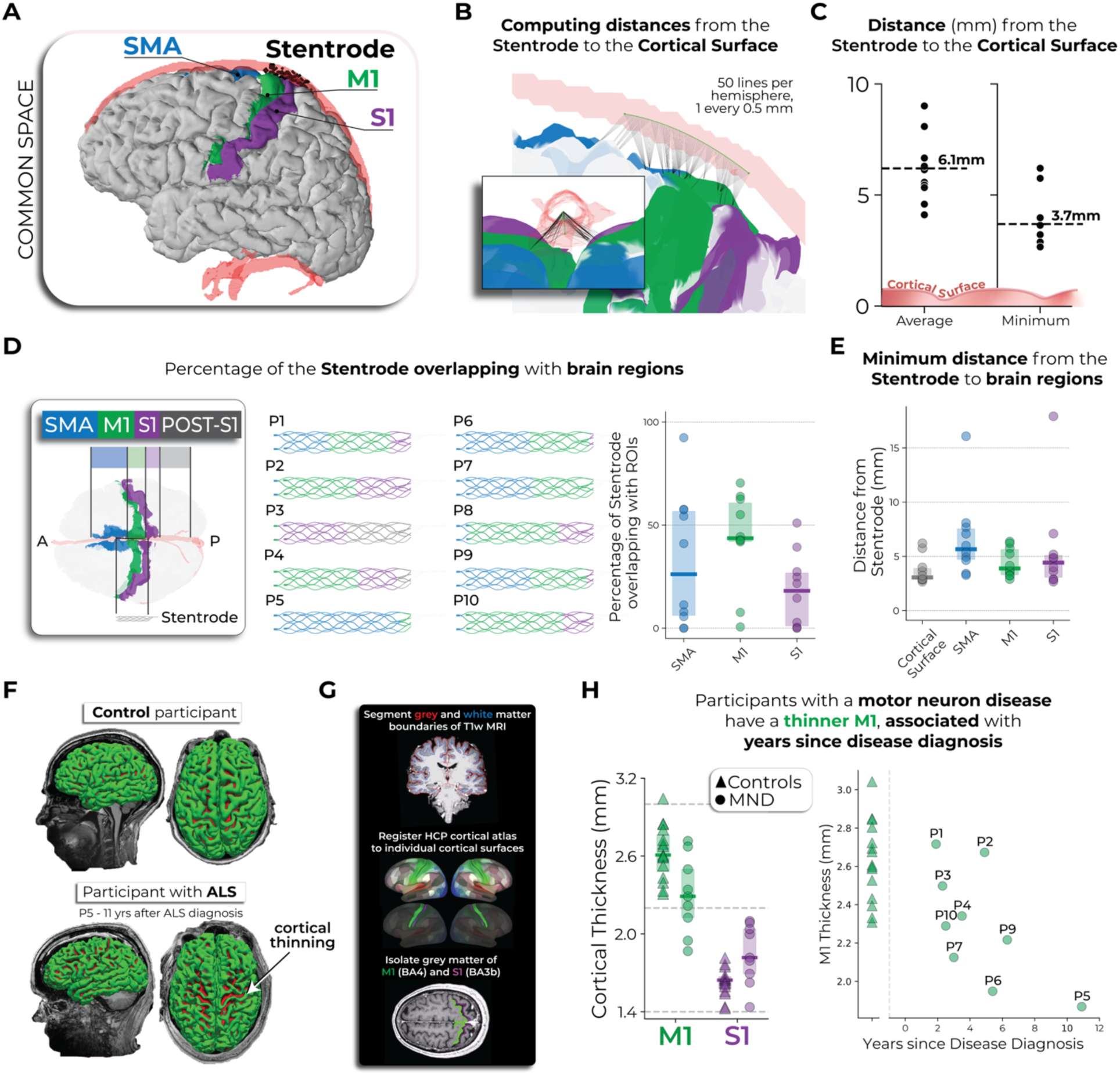
Peri-implant neuroanatomical environment of Stentrode-implanted participants. **(A)** An example participant’s pial cortical surface visualized with their Stentrode segmentation, SSS segmentation, and relevant cortical regions (SMA, M1, S1) registered to the cortical surface. Blue=SMA; green=M1; purple=S1. **(B)** Approximation of the Stentrode implant position in the SSS and its distance to the cortical surface. Black lines indicate shortest distances to cortex. **(C)** The average (data on left) and minimum (data on right) distance from the Stentrode line to the cortical surface is displayed. **(D-E)** Spatial overlap and proximity of each participant’s Stentrode with SMA, M1 and S1. **(F)** Grey matter of a non-ALS control participant (top) and Stentrode-implanted participant with ALS (bottom; 11 years after ALS diagnosis). **(G)** Method for quantifying cortical thickness. **(H) Left** – Stentrode-implanted participants with motor neuron diseases (MND; shown as circles) exhibited a thinner M1, not S1, relative to a group of control participants (shown as triangles). **Right** – M1 cortical thickness was associated with the number of years since each participant’s disease diagnosis, i.e., M1 thins with motor neuron disease progression.

We next examined the extent to which the Stentrode overlapped with specific brain regions: the SMA (blue), M1 (green), S1 (purple), and any territory caudal to S1 (grey; Figure 4D). While each participant had a unique distribution of overlap across these regions, M1 was the predominant area of coverage for most participants, except for P3 and P5 (for a visualization of all participants cortical surfaces and Stentrode implant position see Figure 3). We repeated the distance calculations shown in Figure 4C for each ROI and found that across participants, the Stentrode was closest to M1 (Figure 4E). Considering this proximity to M1, one important consideration is each participant’s disease progression, since motor neuron disease pathology triggers motor neuron death and M1 cortical atrophy^21–27^. M1 thinning might increase the distance between the Stentrode and desired cortical motor signals, potentially weakening signal strength. For example, see the cortical anatomy of the most progressed participant with ALS in our cohort (P5: 11 years after diagnosis), relative to a control participant without ALS (Figure 4F). Data from 16 control participants of a similar age were pooled from a pre-existing dataset^28^ (age of participants with motor neuron diseases vs. control participants: Mann-Whitney independent samples test: W=109.0, *p*=0.13). To assess whether cortical thinning was observed among Stentrode-implanted participants with motor neuron diseases, we segmented the grey matter of M1 and S1 for each participant and computed the average thickness of each (Figure 4G). We observed that participants with motor neuron diseases (P8 excluded) exhibited a thinner M1, but not S1, relative to control participants [n=16; Figure 4H; rmANOVA: ROI*group interaction: F_(1,21)=_19.6,*p*<0.001; Mann Whitney U tests: M1: W=101, *p_corr_*=0.03; S1: W=40, *p_corr_*=0.318]. Further, M1 thickness was significantly associated with the number of years since their motor neuron disease diagnosis, i.e., participants that were more progressed exhibited a thinner M1 (Spearman correlation: *r_s_*=-0.70, *p*=0.036). Further, muscle strength scores were also significantly associated with the cortical thickness of M1 (*r_s_*=0.76, *p*=0.02), and not S1 (*r_s_*=0.13, *p*=0.74), such that participants with less residual muscle strength also exhibited a thinner M1. Combined, the analyses highlight the inter-participant variability in the peri-implant neuroanatomical environment that could potentially contribute to Stentrode signal strength.

### Device integrity

Variability in the number of active recording channels across participants

The Stentrode BCI has 16 recording channels that are connected to a intravascular lead that runs through the vasculature, from the SSS to the internal jugular vein. Following Stentrode deployment, the lead was inserted into an inductively powered internal telemetry unit (ITU*; Synchron, USA*) positioned in the chest. Due to the difficulty of ensuring a perfect fit, this procedure can lead to some channels having an inactive connection. As such, across participants, the number of active recording channels varies (9 to 16), which could potentially impact Stentrode motor signal strength, particularly if the inactive electrodes were those that would have recorded the most selective movement-related information.

Due to a technical challenge—and because the participant was the cohort’s only stroke participant, whereas all other participants had ALS or PLS—we excluded P8 from analyses comparing the user-specific factors to Stentrode motor signal strength.

### Stentrode motor signal strength

Detecting attempted movement using the Stentrode BCI

With multiple user-specific factors computed, we next aimed to compute a single measure of Stentrode motor signal strength for each participant. Crucially, we opted to compute a signal strength measure, as opposed to a BCI performance measure due to changes in decoders and electrode referencing across participants and clinical trials. To compute this measure, participants routinely performed motor signal testing (see ref^14^), where participants were visually-cued to attempt either hand or foot movements to generate a BCI click within 10 seconds, followed by resting (no movement) for 10 seconds (10 repetitions per task run; Figure 5A). Considering this test was performed within the context of the larger clinical trials and procedures steadily evolved over time, it is worth noting the Motor Signal Test was not administered to all participants in the same manner. These inconsistencies introduce variability in the data collected, including differences in the number of blocks performed per session, total sessions per participant, and the type of imagery strategy attempted (hand or ankle; unilateral or bilateral). Though, to be consistent across participants, we focused on datasets when participant’s used their preferred imagery strategy (n=3 hand-based; n=7 ankle-based; Supp. Table 1; for data of all imagery strategies see Supp. Figure 4). For each test block, we isolated the high-gamma frequency band (100 – 200 Hz) across all Stentrode channel recordings and computed a sensitivity index between the density of the burst count during the move epochs versus the rest epochs (see Methods; Figure 5B). Hereafter, we refer to this sensitivity index measure as Stentrode motor signal strength^14^.

**Table 1.** Participant demographics. Regarding the host country of the clinical trial, the SWITCH trial was conducted in Australia (AUS) and the COMMAND EFS trial was conducted in the USA. Y=yes; N=no; PLS=primary lateral sclerosis; ALS=amyotrophic lateral sclerosis.

| **Participant ID** | **P1** | **P2** | **P3** | **P4** | **P5** | **P6** | **P7** | **P8** | **P9** | **P10** |
| --- | --- | --- | --- | --- | --- | --- | --- | --- | --- | --- |
| **Sex** | M | M | M | M | M | M | M | F | F | M |
| **Disease/injury** | ALS | ALS | ALS | PLS | ALS | ALS | ALS | Pontine arterial ischemic stroke | ALS | Main-in-the-barrel ALS |
| **Years since diagnosis, at time of consent** | 1.9y | 4.9y | 2.3y | 3.5y | 10.9y | 5.4y | 3.0y | 13.5y | 6.3y | 2.5y |
| **Mechanically ventilated, during study** | N | N | N | N | Y | Y | Y | N | N | N |
| **Preferred imagery strategy for Stentrode control** | Left ankle | Both ankles | Right ankle | Both ankles | Both ankles | Right hand | Both ankles | Both ankles | Right hand | Right hand |
| **Stentrode device version; # of connected (active) channels** | V1;16 | V1;15 | V1;15 | V1; 15 | V2;15 | V2;9 | V2;13 | V2;13 | V2;12 | V2;10 |

**Figure 5.**
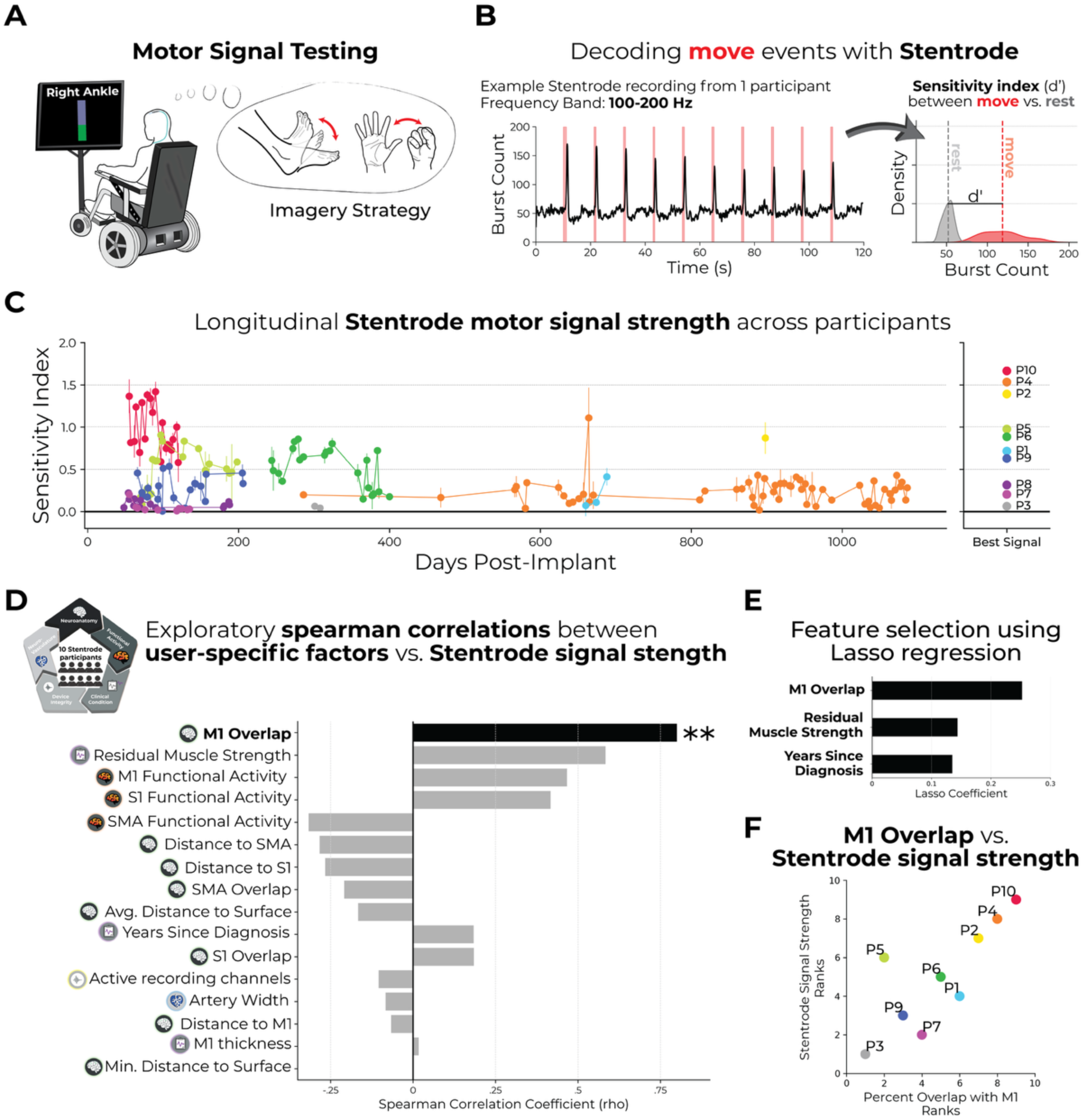
M1 overlap associated with Stentrode BCI motor signal strength. **(A)** To quantify Stentrode BCI signal strength for each participant, participants were visually cued to perform 2 different motor imagery strategies: either an ankle flexion/extension or hand close/open movement (unilateral or bilateral). **(B) Left** – Example Stentrode BCI recording (100-200 Hz frequency band) for 1 participant for a single task session. The red line denotes the on/off period of the movement cue. The black line depicts the Stentrode BCI burst count during the task. **Right** – A signal sensitivity index (d-prime) was computed by comparing burst count densities of ‘rest’ epoched data (shown in grey) versus ‘move’ epoched data (shown in red). **(C) Left** – Longitudinal Stentrode BCI signal strength (i.e., the sensitivity index) values are shown for all participants for all testing sessions. The datasets reflect only the blocks where participants used their preferred imagery strategy. The error bars reflect instances where the task was performed multiple times in a single session. **Right** – The highest Stentrode BCI signal strength value for a single session of each participant are plotted. Participants with the highest session signal strength values are coloured in warm colors and lower in cool colors. **(D)** When correlating Stentrode BCI signal strength to each of the user-specific factors, one significant predictor emerged: percentage of the Stentrode BCI overlapping with M1. **(E)** Similarly, a Lasso regression analysis revealed 3 factors with non-zero lasso coefficients: M1 overlap, residual muscle strength and years since diagnosis. **(F)** Visualization of the correlation between the percentage of the Stentrode BCI overlapping with M1 and motor signal strength (r_s_=0.80, p_uncorr_=0.01; ranked values shown in correlation).

For each participant across testing sessions, Stentrode motor signal strength values were significantly greater than 0 (one Wilcoxon signed-rank test per participant; 0.0001 < *p* < 0.01; P3 was not included in the group analysis due to only having 2 sessions of this task, though both were greater than 0), where a value of 0 reflects no difference in Stentrode recordings during move events and rest events. Across participants and sessions, we observe variability in the magnitude of Stentrode motor signal strength (Figure 5C). To generate a single value for each participant’s Stentrode motor signal strength to compare to the user-specific factors, we opted to select each participant’s best signal strength value captured in any given test block. The rationale for this decision was to mitigate the inconsistencies in how the task was administered between clinical trials and the progressive nature of motor neuron disease. We note that P4’s best session appears to be an outlier relative to the other sessions (Figure 5C), however P4 had similarly high values in other sessions when using their non-preferred imagery strategy (Supp. Figure 4).

### Statistical comparisons

M1 overlap best predicts Stentrode motor signal strength

Using this measure of Stentrode motor signal strength, we next performed exploratory Spearman correlations to the multiple user-specific factors extracted (16 tests total). Across all tests, we identified only one significant predictor: the percentage of the Stentrode overlapping with M1 (*r_s_*=0.80, *p_uncorr_*=0.01), such that greater M1 overlap reflects increased Stentrode motor signal strength (Figure 5D; for a visualization of all ranked correlations see Supp. Figure 5). To explore the relationships between Stentrode signal strength and the user-specific factors more rigorously, we also performed a feature selection using Lasso regression. This analysis allowed us to identify which factors were most predictive of Stentrode signal strength while penalizing irrelevant or redundant predictors. Among all factors considered, three factors survived, with greater than zero Lasso coefficients: M1 overlap (the highest), followed by residual muscle strength, and years since diagnosis (Figure 5E). Collectively, these tests show the amount of the Stentrode overlapping with motor cortex is the strongest predictor of Stentrode motor signal strength with clinical measures such as residual muscle strength and disease duration contributing additional, complementary explanatory power when considered in combination. (Figure 5F).

### Considerations for future Stentrode BCI deployment

The previous analyses (Figure 5F) show that M1 overlap is associated with Stentrode BCI signal strength. For future Stentrode implant targeting, this result suggests that deployment should specifically target primary motor cortex, defined by its neuroanatomical landmarks, e.g., the rostral wall of the central sulcus, to maximize Stentrode BCI signal outcomes. However, it is still unclear whether pre-implant fMRI activity can be used to further optimize positioning (Figure 6A). To test this, we aimed to directly compare volume-based fMRI activity versus M1 overlap. We performed a volume-based fMRI activity analysis, where we projected the average pre-implant fMRI motor activity for every slice across the dorsal stream, i.e., the cortical surface directly beneath the SSS vasculature. To select the ideal implant location based on the fMRI activity, we computed the center of gravity of this projection. We next identified, for each participant, the slices that overlap with their Stentrode BCI and the M1 brain region. Finally, we computed the distance between the ideal fMRI slice (i.e., center of gravity of movement-related activity) and the slice at the midpoint of where the Stentrode BCI was implanted (Figure 6B). Often the centroid of activity was rostral to the midpoint of the Stentrode. For five out of ten participants, the centroid of activity was within M1; for four participants the centroid was rostral to M1 and for one participant it was caudal to M1. We also computed the percentage of slices that encompassed the Stentrode BCI that overlapped with M1 (Figure 6B). Similar to the surface-based analysis, we observed that M1 overlap, defined on the volume vs. on the surface, also showed a significant association with Stentrode BCI signal strength (Spearman correlation: *r_s_*=0.70, *p*=0.043). Alternatively, there was no meaningful relationship between Stentrode signal strength and how far away the Stentrode BCI was from the location of peak fMRI activity (*r_s_*=-0.18, *p*=0.64; Figure 6C). Combined, with the previous analyses, this confirms future Stentrode BCI deployment should prioritize maximizing motor cortex coverage and the present functional MR imaging protocol may be insufficient for improving targeting.

**Figure 6.**
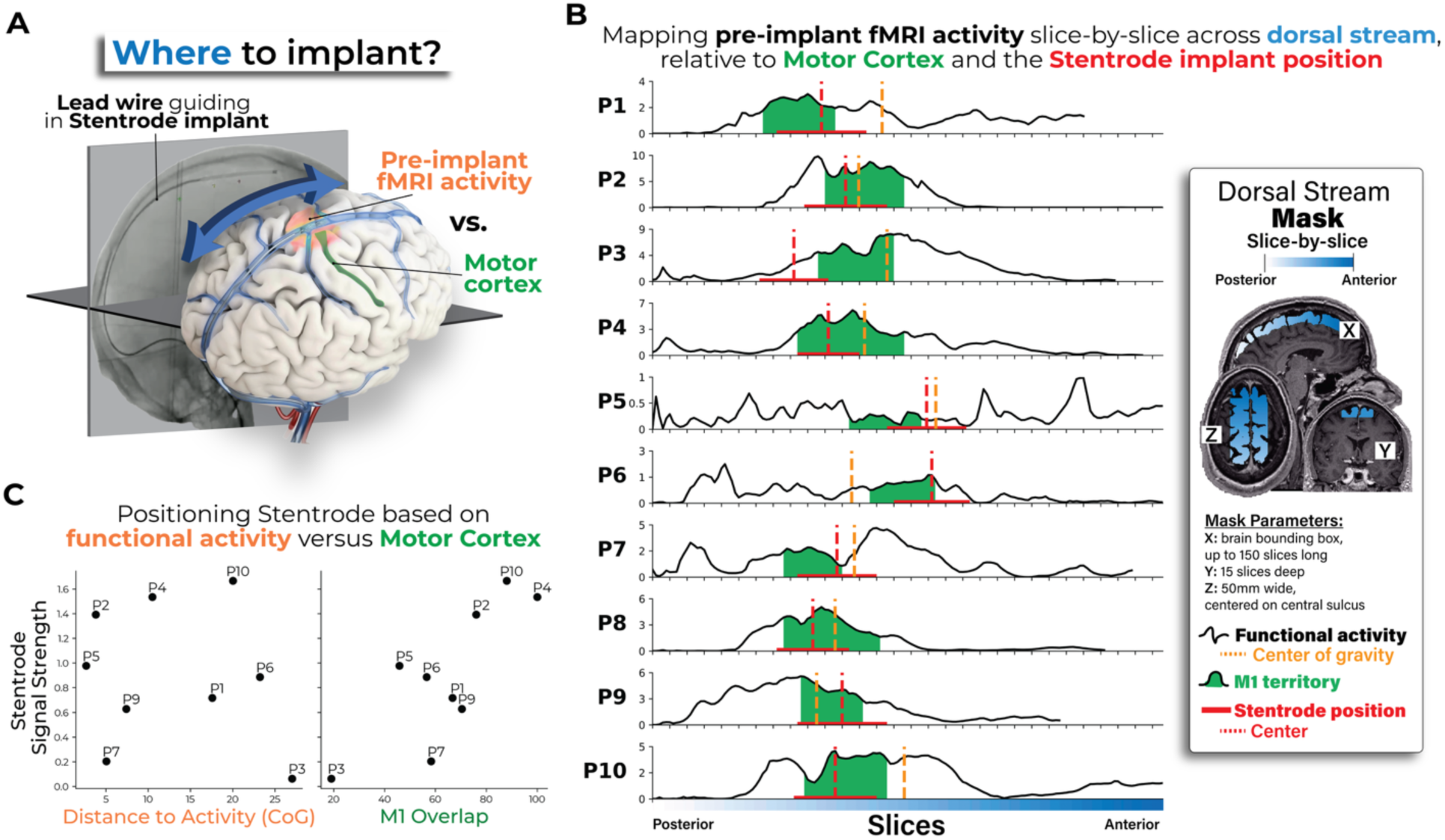
Motor cortex coverage, over functional activity, is strongly associated with Stentrode BCI signal strength. **(A)** An important consideration when deploying the Stentrode BCI via a lead wire is deciding where to position the device within the superior sagittal sinus. There are two factors to base this decision on: (1) pre-implant fMRI activity, relying on the location of the largest cluster of activity or (2) neuroanatomical landmarks, i.e., motor cortex on the rostral wall of the central sulcus. **(B)** To test which factor impacted signal strength in the participants, we performed a volume-based analysis. For each participant (each row), we projected the average pre-implant fMRI motor activity (black line) for each slice across the dorsal stream (white-to-blue colored mask). We then computed the center of gravity of this projection across the dorsal stream, depicted as a yellow dashed vertical line. This would reflect the best implant site if based solely on the fMRI activity. We next defined the slices that overlap with the M1 cortical region (colored in green) and the slices that overlap with the Stentrode BCI (red horizontal line; the center point depicted as a red dashed vertical line). **(C)** Finally, for each participant, we then computed (1) the distance between the fMRI center of gravity slice to the Stentrode BCI center slice and (2) the percentage of Stentrode slices overlapping with M1 slices, i.e., M1 coverage. We observed that M1 coverage showed a significant association with Stentrode signal strength (Spearman correlation r_s_=0.70, p=0.043). Alternatively, there was no meaningful relationship with the distance to the ideal fMRI slice and Stentrode BCI signal strength (r_s_=-0.18, p=0.64).

## Discussion

Across 10 participants implanted with a Stentrode BCI, we explored the relationship between a variety of user-specific factors and Stentrode BCI motor signal strength. Regardless of impairment level or diagnosis (ALS, PLS or brainstem stroke), participants were able to functionally activate cortical motor networks during attempted movement prior to implantation, confirming that the ability to voluntarily produce cortical motor signals were preserved in the cohort, at least to some degree. Structurally, we observed that the study participants with motor neuron diseases exhibited motor cortical atrophy relative to controls, with cortical thickness correlating with both participants’ residual muscle strength and time since disease diagnosis.

Additionally, we quantified the inter-participant variability of the peri-implant neural environment across all participants implanted with a Stentrode BCI, relative to specific cortical regions and the neurovasculature. Most importantly, across all tested user-specific factors, we identified that the strongest predictor of BCI motor signal strength was the degree to which the Stentrode implant overlapped with primary motor cortex (M1), emphasizing the importance of precisely targeting M1 during device deployment. Our findings have three main implications for clinical neuroscience research and the development of BCI technologies. Specifically, it (1) highlights the unique neural and clinical considerations for BCI signal strength for individuals living with adult-onset motor neuron diseases, (2) informs the future targeting strategy for Stentrode BCI deployment (and other future BCIs that may use an endovascular electrode deployment approach) and, (3) offers a general investigational framework for testing the impact of user-specific factors on signal strength of BCI technologies.

First, our results offer a high-level characterization of the neural and clinical factors that need to be considered for successful BCI usability in individuals living with adult-onset motor neuron diseases, a primary target clinical population for BCI technologies. Most common in our cohort, ALS is a heterogenous neurodegenerative disease, marked by progressive motor decline and generally poor prognosis; population studies report a median survival of ∼3 years from symptom onset in unventilated cohorts^29^, but survival can extend well beyond this in individuals who opt for long-term mechanical ventilation or present with slower-progressing variants, such as the flail-arm phenotype that characterized many participants in the present cohort. For people with ALS to successfully use a Stentrode BCI—or any BCI—it is essential they have some intact structure and function of cortical motor networks, with M1 being the dominant region of interest. However, decades of research has investigated how ALS pathology uniquely degrades M1 micro- and macro-structure and functioning. ALS pathology most commonly first targets lower motor neurons, followed by pathological changes throughout cortex, particularly a degradation of the upper motor neurons in M1^19,20,30^. Though, the symptomatic presentation of ALS is highly heterogeneous. Indeed, M1 is the key hub for voluntary motor command output, and multiple studies have demonstrated its vulnerability in later stages of ALS. For example, post-mortem and high-field neuroimaging studies have highlighted that individual’s with late-stage ALS exhibit multiple pathologies to M1 micro-cortical structure, including the degeneration of Betz pyramidal cells in layer V^30^, the accumulation of iron in deeper cortical layers^19,23^, increased intracellular calcium^31^, and progressive, widespread demyelination, particularly at the boundaries between somatotopic regions^19^. In terms of M1 macrostructure, similar to our findings, several cross-sectional studies have demonstrated that individuals with ALS exhibit dramatic cortical atrophy of the precentral gyrus (i.e., pre- and primary-motor cortex), compared to individuals without ALS^21–27^. For example, in a recent report of a long-term ECoG BCI case study, longitudinal CT imaging data from 1 participant revealed significant fronto-temporal cortical tissue atrophy over an 8-year period^32^. Notably, for our investigation, neither M1 cortical atrophy nor the minimum distance between the Stentrode and motor cortex were significant predictors of Stentrode BCI motor signal strength (Figure 5D). Instead, the significant association we observed was with the percentage of the Stentrode BCI overlapping with M1–a distance measure solely on the coronal plane–suggesting that larger Stentrode BCI motor signal strength may be attributed to the maximum number of Stentrode electrodes being closest to motor cortex versus other cortical regions. Indeed, even though the motor cortex progressively atrophies with disease progression, any space between motor cortex and the vasculature is filled with an expanding gap of cerebrospinal fluid (CSF; Supp. Figure 3). Our results suggest that so long as the overlap with M1 is sufficient, the high electrical signal conductivity of cerebrospinal fluid^33^ may still effectively be delivering a selective motor signal to the Stentrode BCI electrodes. This would suggest that cortical atrophy alone should not be an exclusion criteria for Stentrode BCI usability, especially given prior BCI studies demonstrating successful motor decoding in individuals with ALS^14,32,34–37^.

Next, looking to M1 cortical function, neuroimaging studies have reported mixed findings on whether individuals with ALS—particularly those who are locked-in—can volitionally activate M1 during attempted movements similar to control participants (see ref.^38^ for a comprehensive review of task-based neuroimaging studies of ALS). While some studies, including a recent high-field neuroimaging case report, observed preserved M1 activity during motor tasks^39^, others have shown highly variable or diminished responses, likely reflecting the extreme heterogeneity of ALS^38^. This preservation in cortical motor activity, at least to some degree, could be explained by evidence that people with late-stage ALS show a considerable amount of preservation of the upper motor neurons of M1 output layer V^40–42^, at least sufficient for generating a motor output signal. However, it remains an open question whether the presence or strength of M1 activity pre-implant can be used as a reliable predictor of BCI performance. The intricate relationship between M1 structure and function and clinical status—including disease duration and residual muscle strength—likely contributes to BCI signal strength. While no single clinical measure was individually significant in explaining variability, both residual muscle strength and years since diagnosis were retained as predictors in the Lasso regression model. This suggests that, although their individual effects may be subtle, these clinical factors may contribute meaningful, non-redundant information when considered alongside the implant’s proximity to M1. These findings underscore that successful BCI use in people with ALS is not determined by any single clinical or neural factor, but instead reflects a complex interplay between disease progression, preserved cortical function, and precise anatomical targeting— highlighting the need for multimodal, individualized assessment in guiding future BCI deployment.

The most practical implication of our results is they provide immediate guidance on future endovascular BCI deployment strategies, specifically to what degree functional neuroimaging data should be relied on as a targeting guide versus neuroanatomical landmarks (e.g., M1 located on the rostral wall of the central sulcus). For both surface- and volume-based analyses, Stentrode BCI overlap with M1 emerged as a predictor of Stentrode BCI motor signal strength. Further, across participants, the spatial landscape of pre-implant functional activity during attempted movement often lacked a clear unimodal structure along the coronal plane, with the largest peak appearing in different regions (i.e., S1, M1 or SMA) at the individual participant level (Figure 6). In order for personalized functional neuroimaging to be a useful targeting guide, spatially selective functional activations are essential to identify an optimal deployment site. However, in order to visualize more spatially selective activations, it requires changing the MR imaging protocol to incorporate (1) more functional data (e.g., more than the 1-2 runs per participant collected in the present study), and (2) more movement conditions to control for regions responsive to general motor responses, the presence of visual/auditory stimuli during the active epochs, and attentional/motivational drivers of activity. While there is a simple solution, increasing scan times for people with motor neuron diseases, particularly those in advanced stages, has considerable tradeoffs. For example, in the present cohort, 3 participants were mechanically ventilated via a tracheostomy and had minimal to no residual volitional movement of a single body-part (see Supp. Table 1 for descriptions of each participants’ clinical state). Scanning these participants required participants being moved off their personal ventilators, manually ventilated by a respiratory therapist and then placed on an MR-compatible ventilator for the duration of the scanning, regularly monitored by a respiratory therapist. During the scan, the respiratory team had to ensure participants had a clear airway, requiring monitoring of the participant’s residual eye movements and physiological recordings (e.g., pulse oximeter, EKG, and the ventilator, etc). Therefore, due to burden of longer functional scan times in these participants, our recommended guidance for Stentrode deployment is to prioritize maximizing coverage of M1, specifically the rostral wall of the central sulcus. To the extent that the Stentrode BCI is larger than M1, it may be beneficial to bias the positioning more rostral so as to be able to record activity from SMA as well, given its putative involvement in higher order motor and cognitive processes that may be beneficial for complex BCI control^43,44^. However, while present device deployment may not warrant necessitating functional neuroimaging for targeting, next-generation devices could be engineered to target smaller cortical vessels, where a smaller cortical target is necessary. In these instances, functional neuroimaging protocols will need to be improved to ensure a sufficient amount of functional data is acquired to guide targeting of smaller cortical sites. More generally, for BCI devices with broad cortical coverage (i.e., the Stentrode BCI, larger ECoG arrays), we recommend prioritizing anatomical targeting— specifically M1—over individualized functional imaging, that is unless neuroimaging protocols can be optimized (e.g., increased scan times and conditions) to yield sufficiently selective activations without imposing undue risk or burden on participants with advanced disease.

There are several limitations with our investigation that are worth noting. First, we did not perform any statistical corrections for the multiple comparisons, e.g., the 16 user-specific factors tested (Figure 5D-E). The rationale for this decision was due to the highly exploratory nature of the investigation. Our hope is that this data serves as a proof-of-concept to guide the design of future studies with larger cohorts, as they become available. A second limitation is that our estimation of the Stentrode BCI position was accurate solely along the coronal plane, but not accurate in relation to the unique locations of individual electrodes within the SSS. Due to the methodological difficulties with reliably extracting single electrode positions from the CT images and computing distances for each electrode to various targets, we opted for a simpler estimation that was accurate along the coronal axis and positioned within the center of the SSS segmentation. A third limitation is that we provide a measure of motor BCI signal quality, not BCI performance. Finally, considering the aim of the investigation was to pool data across 2 clinical trials (spanning over 5 years), there are differences in the referencing scheme of the Stentrode BCI and the experimental design of the Motor Signal Test used to estimate Stentrode BCI motor signal strength between the two study cohorts. Despite these differences, we did not observe any differences in Stentrode BCI motor signal strength between participants from the two trials, with a similar percentage of participants from each trial as high-responders (upper-half of the sensitivity index distribution) and low-responders (lower-half of the sensitivity index distribution; independent samples Mann-Whiteny U test; W=13.0; *p*=0.914).

In conclusion, our study provides a comprehensive scientific framework for investigating how a variety of user-specific factors contribute to recorded signal strength of implantable BCI technologies. As more individuals are implanted with BCI devices, identifying key user-specific predictors of BCI success will be essential for the long-term viability of the neurotechnology industry. These predictors will be vital for refining patient selection, optimizing implant targeting strategies and tailoring BCI hardware and software for specific clinical populations. By identifying potential determinants of successful neural interfacing with the Stentrode BCI, we hope this investigation offers some foundational insights into improving the clinical translation of the Stentrode BCI and, more broadly, contribute to the advancement of all implantable BCI technologies.

## Supplementary Results

**Supp. Figure 1.**
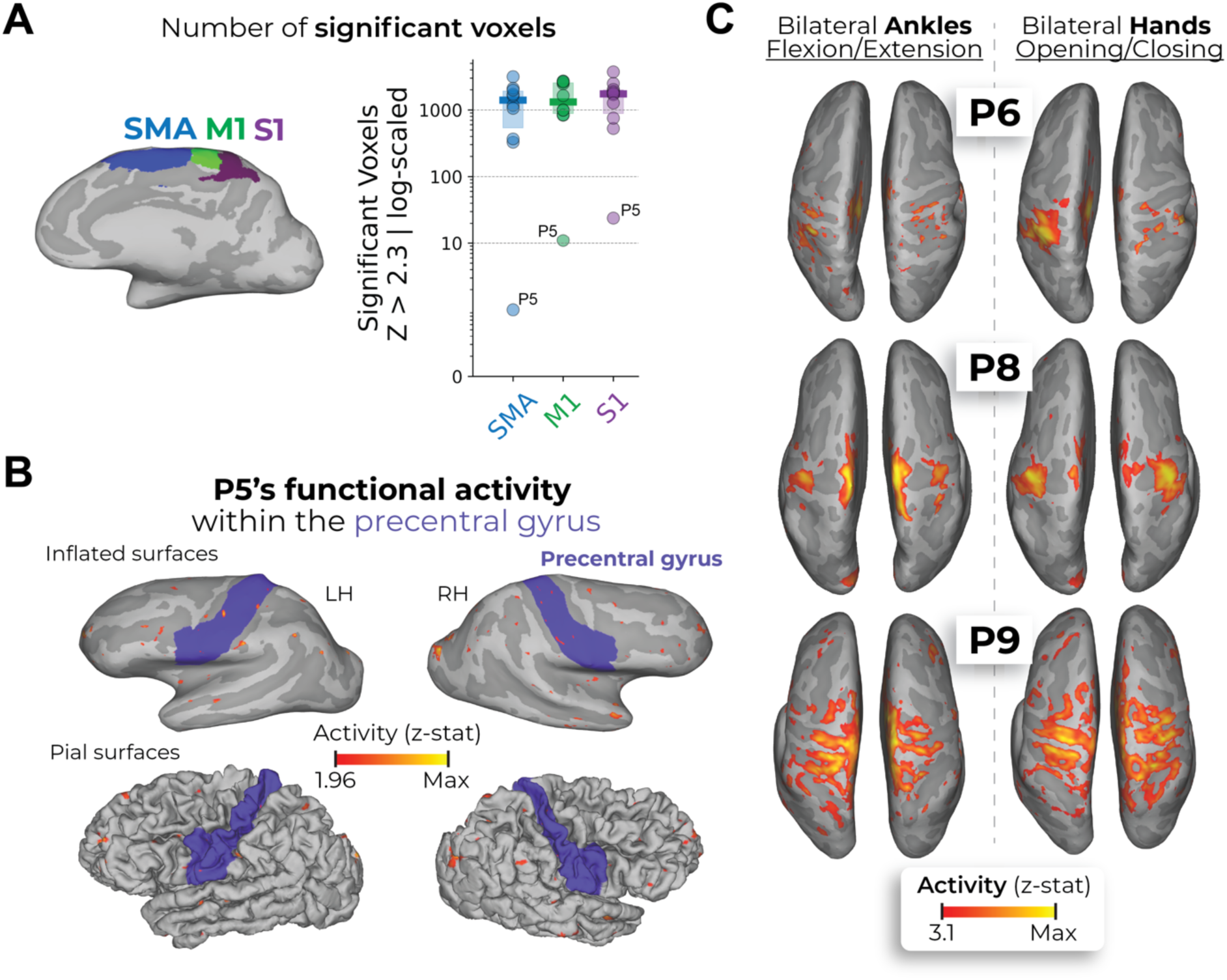
Supplementary functional neuroimaging analyses. **(A)** Number of significant voxels (z-statistic > 2.3; z-threshold used in Figure 2) within the SMA (blue), M1 (green) and S1 (purple) regions. All participants can significantly activate a subset of voxels, within all regions, during attempted movement. The data was visualized on a log-scale to demonstrate that P5 significantly activated a subset of voxels. **(B)** Visualizing P5’s functional data at a lower minimum threshold (z-statistic > 1.96) within the precentral gyrus highlighted (in blue) on both the inflated surfaces (top row) and pial surfaces (bottom row). **(C)** A subset of participants underwent functional scans where bilateral hand movements were performed. The bilateral ankle and bilateral hand activation maps are displayed. All other annotations are the same as described in Figure 2.

**Supp. Figure 2.**
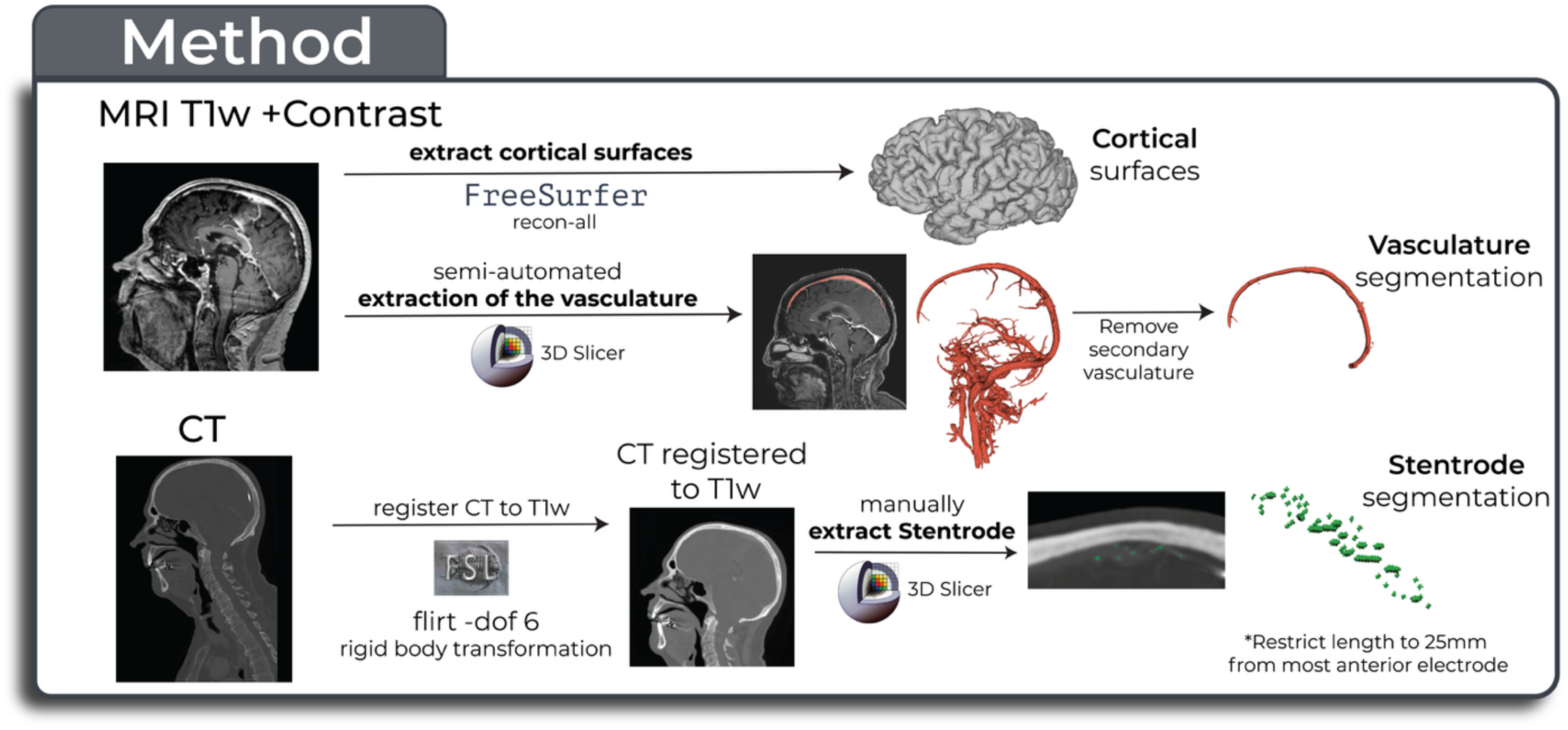
Method for generating a common coordinate space with the cortical surfaces, vasculature and Stentrode BCI. See the Methods for a detailed description of the method.

**Supp. Figure 3.**
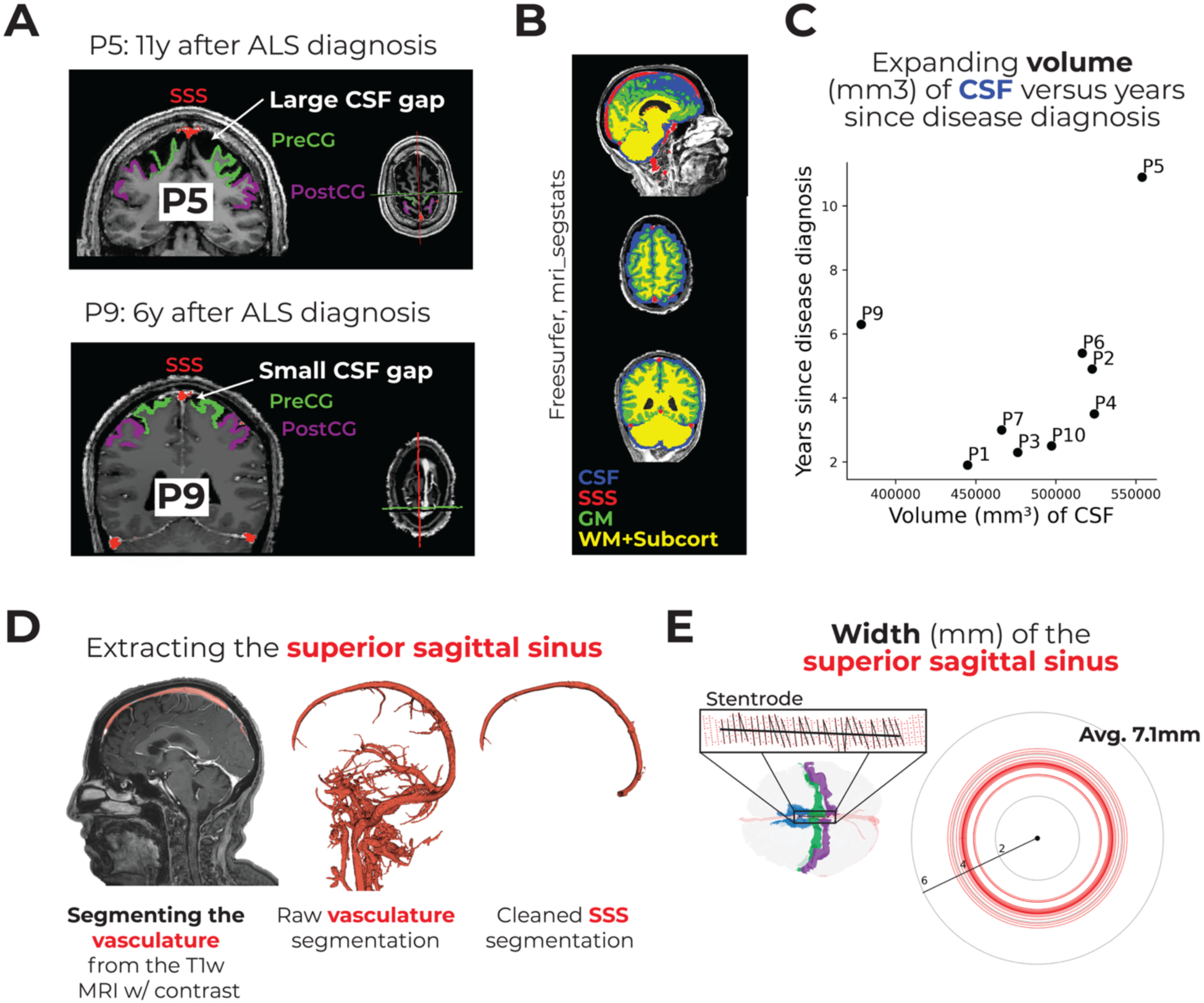
Segmenting the cerebrospinal fluid (CSF) and superior sagittal sinus (SSS) for Stentrode-implanted participants with adult-onset motor neuron diseases. **(A)** Visualizing the SSS, pre- and post-central gyri on the T1w + contrast MRI for the two participants with the most progressed ALS: P5 (top row) and P9 (bottom row). P5 shows an extensive gap, in this extreme slice example, between the SSS and the cortical surface filled with CSF. Alternatively, P9 generally shows a much more minimal gap. **(B)** To segment the CSF, we used freesurfer’s mri_segstats function to generate segmentations of the CSF (blue), grey matter (GM; green), white matter and subcortical structures (both shown in yellow). As the CSF segmentation includes the SSS, we removed any SSS segmentation voxels from the CSF segmentation. We then computed a measure of whole-brain CSF volume (mm^3^) from the segmentation. **(C)** We observed a significant association between years since motor neuron disease diagnosis and total volume of CSF (r_s_=0.83, p=0.01), though not when P9 was included (r_s_=0.41, p=0.2). **(D)** A visualization of one participant’s segmentation of the neurovasculture overlaid on the T1w MRI, the raw vasculature segmentation in 3D space and the manually cleaned segmentation of the SSS. **(E)** Using the SSS segmentation, the diameter of the SSS was measured to be on average 7.1 ± 0.9 mm across Stentrode implanted participants. Individual red circles reflect the average SSS width for the points where the Stentrode is implanted.

**Supp. Figure 4.**
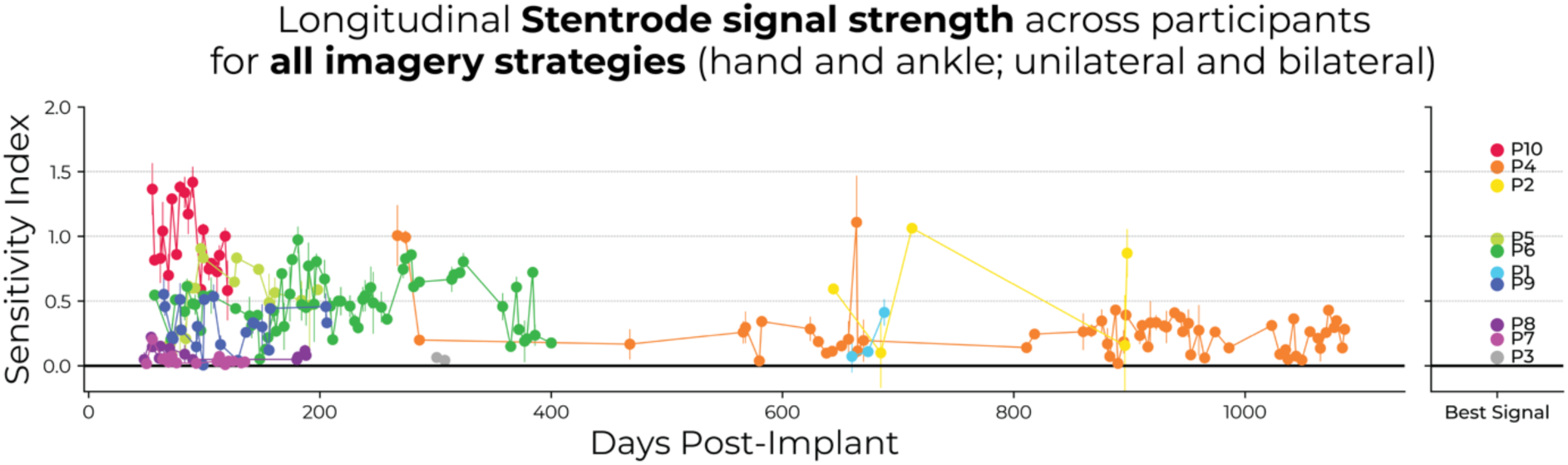
Stentrode motor signal strength datasets for all imagery strategies. While the data plotted in Figure 5C is only for the preferred imagery strategy of each participant (either hand or ankle), the data displayed here includes all imagery strategies.

**Supp. Figure 5.**
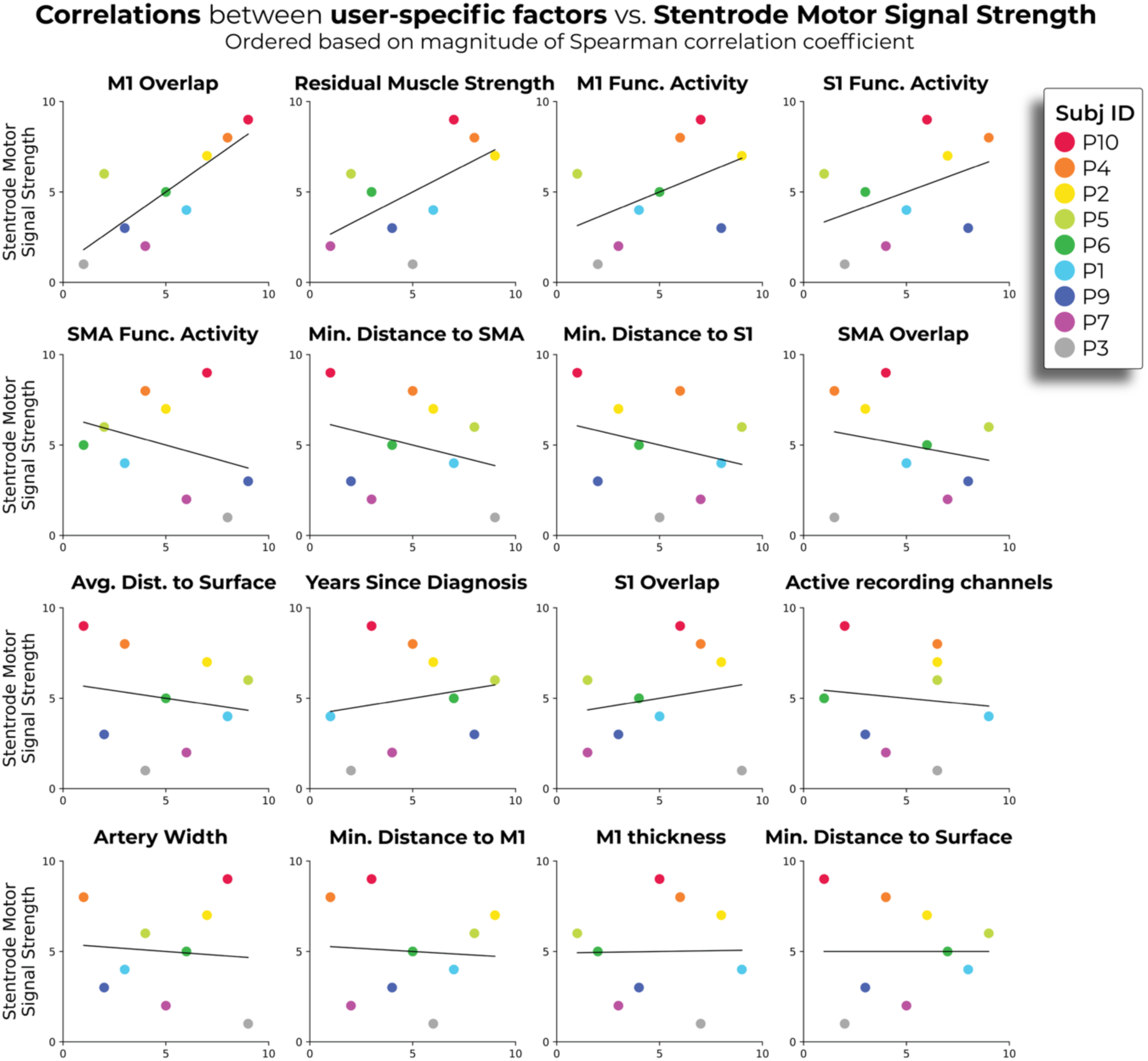
Plotting correlations between all user-specific factors and Stentrode motor signal strength. All plotted data is the ranked values.

## Methods

### Clinical trials and participant recruitment

Study participants were involved in 1 of 2 clinical trials: SWITCH or COMMAND. The SWITCH trial (n=4; participants P1 – P4; Clinicaltrials.gov: NCT03834857) was approved by the Human Research Ethics Committees of St. Vincent’s Hospital, Royal Melbourne Hospital and Calvary Health Care Bethlehem in Australia. The clinical protocol for this trial has been previously published^10,11^. The COMMAND early feasibility trial (n=6; participants P4 – P10; Clinicaltrials.gov: NCT05035823) was conducted under an Investigational Device Exemption from the U.S. Food and Drug Administration and approved by the Institutional Review Boards at Western-Copernicus Group (1347924). Informed consent was obtained before any study procedures were conducted and in accordance with the Declaration of Helsinki.

### Stentrode device and deployment

The Stentrode (Synchron, USA) consists of 16 platinum electrodes, each with a 300 *μ*m diameter, mounted on a self-expanding nitinol scaffold measuring 8 × 40 mm. The inter-elctrode spacing is approximately 3mm. The Stentrode is connected to a 50cm flexible endovascular lead and inserted into an inductively powered internal telemetry unit^11^ (ITU; Synchron, USA). The device, surgical deployment procedure and signal acquisition have been previously described in detail^10,11^. In the SWITCH trial, a common reference channel was fixed to an electrode located on the ITU. In the COMMAND trial, the reference channel was configurable across one of the 16 channels or the electrode on the IRTU.

### Participants

#### Stentrode BCI group

The Stentrode BCI group included ten participants [two females; mean age ± STD = 58.4 ± 12.6]. Eight participants were diagnosed with ALS, one participant diagnosed with PLS (P4), and one participant diagnosed with an arterial ischemic stroke in the brainstem (P8; see Table 1 and Supp. Table 1 for all participant demographics).

**Supplementary Table 1.** Qualitative description of the functional status of participants.

| **Participant ID** | **Baseline Functional Status** |
| --- | --- |
| P1 | Limited mobility of upper and lower body starting out, eventually developed pseudo-bulbar issues (e.g., intense coughing). |
| P2 | Slow progressive condition; upper body started losing functionality and very slowly affected his lower limbs, causing him to be wheelchair bound. Breathing difficulties made it difficult for him to speak and made him tire easily. |
| P3 | Fast progressing condition; affected his whole body. |
| P4 | Slow progressing condition; limited movement of lower limb, but proficient upper limb movement. Had an injured right shoulder making him unable to raise hand above shoulder level. |
| P5 | Locked in; mechanically ventilated via a tracheostomy to breathe, only able to slightly move left side of mouth to indicate 'no' and looks up to indicate 'yes'. |
| P6 | Locked in; mechanically ventilated via a tracheostomy to breathe, only able to slightly smile and very slight movement of right index finger. |
| P7 | Locked in; mechanically ventilated via a tracheostomy to breathe, no residual movement. |
| P8 | Room air via a tracheostomy to breathe; can make noises with passy-muir valve; residual movement in left hand; looks up and down to indicate 'yes' and 'no' |
| P9 | Used supplemental oxygen and a gastrostomy feeding tube; could blink and slightly smile; could make slight noises to indicate distress |
| P10 | Man-in-the-barrel ALS; minimal control of upper limbs; ambulatory and verbal. |

#### Control group

The control group included 16 participants [9 females; mean age ± STD = 53.1 ± 6.37; all right-handed]. The only datasets analyzed from these participants were their structural T1w MRI scans (acquired from a publically available dataset; see ref.^28^ for additional information about these participants, including MR scanning procedures). Importantly, there were no significant differences between the Stentrode BCI group (with motor neuron diseases) and the control group in age (Mann-Whitney independent samples test: W=109.0, *p*=0.13) or sex [though trending; Chi-Squared test: *X_2_*(1)=3.3, p=0.06].

### Manual muscle strength testing

Prior to implantation, participants underwent manual muscle strength testing. This involved a neurologist or physiatrist grading muscular contractions for different movements. For SWITCH trial participants, the following movements were assessed: fingers, wrist, elbow, shoulder, hip, knee, ankle, plantar flexion and dorsiflexion. For COMMAND trial participants, the following movements were assessed: finger abduction (pinky), middle finger flexion, wrist extension, elbow flexion, elbow extension, hip flexion, knee extension, ankle dorsiflexion, ankle plantar flexion and big toe dorsiflexion. For all participants, movements were assessed separately for the left and right sides of the body using the MRC muscle power scale^16,17^, ranging from 0 to 5. The scores reflect the following: 0 = no muscular contraction; 1 = visible muscle contraction, but no movement at joint; 2 = movement at the joint, but not against gravity; 3 = movement against gravity, but not against resistance; 4 = movement against resistance, but less than full; 5 = movement against full resistance, normal strength. From this data, a bilateral whole-body compound score was generated. Scores were averaged for each body-part across sides (left, right) first and then averaged across body-parts, resulting in a measure of manual muscle strength, which we refer to as residual muscle strength.

### Pre-implant MRI overview

Prior to implantation, all participants underwent an MRI session which included structural and functional imaging. Relevant for the present investigation, all participants underwent functional motor mapping scans and a T1w structural scan with a contrast agent.

### Functional motor mapping

To assess whether participants could activate cortical motor networks during attempted movement, all participants underwent functional MRI scans while attempting to perform one or more movements of individual body-parts including: right ankle, left ankle, both ankles or both hands. For ankle movements, participants were instructed to repeatedly flex and extend their ankles. For hand movements, participants were instructed to repeatedly open and close their fingers. Importantly, the body-parts tested varied across participants, due to site-specific modifications to the imaging protocol catered to each participant. To guide participants’ movements during scanning, instructions were delivered using audio-(i.e., Avotec MR-compatible headphones) and/or text-cues, displayed via a visual display projected into the scanner bore. The method for delivering cues (audio, text or both) varied across participants, due to varying technical capacities at each scan site.

The functional task design was structured in a block-design, where each block consisted of a 15s OFF (“REST”) period followed by a 15s ON (“MOVE”) period. The number of block repetitions varied between 8 – 10 across participants. The following participants performed 8 blocks: P1, P2, P3, P4, P6, P7, P10, followed by a 15s rest period. P8 performed 9 blocks followed by a 15s rest period. P5 and P9 performed 10 blocks. Finally, the number of functional runs varied from 1 – 4 depending on participants. For a detailed breakdown of the body-parts and number of functional runs tested per participant, see Supp. Table 2 below.

**Supplementary Table 2.** MRI scanning information for participants. L=left; R=right; FA=flip angle; FOV=field of view; TR=repetition time; TE=echo time; TI=inversion time; “**…**” = same as above.

| ID | MR Scanner;<br>RF Coil;<br>Site Location | Structural MRI (T1<br>MPRAGE) | Functional MRI (BOLD) | Body-parts tested<br>(# of functional runs;<br># of motion outlier TRs > .9mm) |
| --- | --- | --- | --- | --- |
| P1 | 3T Siemens Prisma<br>32Ch<br>Melbourne, AUS | TR=1900ms<br>TE=2.34ms<br>TI=900ms<br>FA=8°<br>VoxelSize=0.9x0.9x1mm<br>FOV=288x288mm<br>Slices=208<br>Slice thickness=1.0mm | TR=2500ms<br>TE=30ms<br>Volumes=85<br>FA=90°<br>VoxelSize=3.75x3.75mm<br>FOV=64 x 64 mm<br>Slices=34<br>Slice thickness=3.0mm | L ankle (1 run; 7)<br>R ankle (1 run; 7) |
| P2 | ... | ... | ... | L ankle (1 run; 0)<br>R ankle (1 run; 0) |
| P3 | ... | ... | ... | L ankle (1 run; 0)<br>R ankle (1 run; 0) |
| P4 | ... | ... | ... | L ankle (1 run; 0)<br>R ankle (1 run; 3) |
| P5 | 3T GE SIGNA Architect<br>32Ch<br>New York, USA | TR=8412ms<br>TE=3.168ms<br>TI=1100ms<br>FA=8°<br>VoxelSize=1mm<sup>3</sup><br>FOV=256x256mm<br>Slices=<br>Slice thickness=1mm | TR=3000ms<br>TE=60ms<br>Volumes=100<br>FA=90°<br>VoxelSize=3.75x3.75mm<br>FOV=64x64mm<br>Slices=<br>Slice thickness=4.0mm | Both ankles (1 run; 0) |
| P6 | 3T GE SIGNA Architect<br>32Ch<br>New York, USA | TR=8412ms<br>TE=3.168ms<br>TI=1100ms<br>FA=8°<br>VoxelSize=1mm<sup>3</sup><br>FOV=256x256mm<br>Slices=194<br>Slice thickness=1mm | TR=3000ms<br>TE=60ms<br>Volumes=85<br>FA=90°<br>VoxelSize=3.75x3.75mm<br>FOV=64x64mm<br>Slices=35<br>Slice thickness=4.0mm | L ankle (1 run; 1)<br>R ankle (1 run; 2)<br>Both ankles (2 runs; run1: 1; run2: 1)<br>Both hands (4 runs; run1: 1; run2: 3; run3: 3; run4: 4) |
| P7 | 3T Siemens Magnetom Biograph mMR<br>32Ch<br>Pittsburgh, USA | TR=1900ms<br>TE=2.34ms<br>TI=900ms<br>FA=8°<br>VoxelSize=0.9mm<sup>3</sup><br>FOV=256x256mm<br>Slices=208<br>Slice thickness=0.9mm | TR=2500ms<br>TE=30ms<br>Volumes=85<br>FA=90°<br>VoxelSize=2mm<sup>3</sup><br>FOV= 110x110mm<br>Slices=32<br>Slice thickness=2.0mm | L ankle (1 run; 0)<br>R ankle (1 run; 0) |
| P8 | 3T Philips Igenia Elition X<br>32Ch<br>Buffalo, USA | TR=7599ms<br>TE=3.4ms<br>TI=900ms<br>FA=8°<br>VoxelSize=0.3x0.3mm<br>FOV=240x240mm<br>Slices=170<br>Slice thickness=1.0mm | TR=3000ms<br>TE=35ms<br>Volumes=95<br>FA=90°<br>VoxelSize=1.79x1.79mm<br>FOV=96x94mm<br>Slices=36<br>Slice thickness=4.0mm | Both hands (1 run; 0)<br>Both ankles (2 runs; run1: 6; run2: 9) |
| P9 | 3T Philips Igenia Elition X<br>32Ch<br>Buffalo, USA | TR=7599ms<br>TE=3.4ms<br>TI=900ms<br>FA=8°<br>VoxelSize=0.3x0.3mm<br>FOV=240x240mm<br>Slices=170<br>Slice thickness=1.0mm | TR=3000ms<br>TE=35ms<br>Volumes=100<br>FA=90°<br>VoxelSize=1.79x1.79mm<br>FOV=96x94mm<br>Slices=36<br>Slice thickness=4.0mm | Both hands (1 run; 6)<br>Both ankles (2 runs; run1: 32; run2: 3) |
| P10 | 3T Siemens Magnetom Biograph mMR<br>32Ch<br>Pittsburgh, USA | TR=1900ms<br>TE=2.34ms<br>TI=900ms<br>FA=8°<br>VoxelSize=0.9mm<sup>3</sup><br>FOV=256x256mm<br>Slices=208<br>Slice thickness=0.9mm | TR=2500ms<br>TE=30ms<br>Volumes=85<br>FA=90°<br>VoxelSize=2mm<sup>3</sup><br>FOV=110x110mm<br>Slices=32<br>Slice thickness=2.0mm | L ankle (1 run; 0)<br>R ankle (1 run; 0) |

### MRI data acquisition

Participants were scanned at 1 of 4 MRI sites based in either: Melbourne (AUS), New York (USA), Buffalo (USA) or Pittsburgh (USA). As such, the scanner, head coil, structural- and functional MRI parameters vary across sites (for a breadown, see Supp. Table 2). MRI images, for all participants, were obtained using a 3-Tesla scanner. Prior to running a structural MR scan, participants underwent a time-resolved angiography with interleaved stochastic trajectories (TWIST) scan whereby a multihance contrast agent was administered. Although the data from this sequence were not used in the current investigation, a subsequent structural anatomical scan was performed using a T1-weighted magnetization prepared rapid acquisition gradient echo sequence (MRPAGE) in the sagittal plane with residual contrast agent from the TWIST angiography. The structural scan sequence parameters vary across participants, described in detail in Supp. Table 2. Next, functional data, based on the blood oxygenation level-dependent (BOLD) signal, were acquired using a gradient echo-planar imaging (EPI) sequence which included slices (32 or 34) with a slice thickness of either 2.0mm or 3.0mm, a repetition time (TR) of either 2500ms or 3000ms, and an echo time (TE) of 30ms. All other scan parameters are described in Supp. Table 2. The motor fMRI scans had either 85, 95 or 100 volumes, depending on the participant (approximately 5 minutes).

### fMRI analysis

Functional MRI data were processed by using FMRIB’s Expert Analysis Tool (FEAT; Version 6.0), part of FSL (FMRIB’s Software Library, www.fmrib.ox.ac.uk/fsl), in combination with custom bash, Python (version 3) and MATLAB scripts (R2019b, v9.7, The Mathworks Inc, Natick, MA). Cortical surface reconstructions were produced using FreeSurfer [v. 7.1.1^45,46^] and Connectome Workbench (humanconnectome.org) software.

### fMRI preprocessing

The following pre-statistical processing was applied: motion correction using MCFLIRT^47^, non-brain removal using BET^48^, spatial smoothing using a Gaussian kernel of FWHM 5mm for the functional task data, grand-mean intensity normalization of the entire 4D dataset by a single multiplicative factor, and high-pass temporal filtering (Gaussian-weighted least-squares straight line fitting, with σ = 100 s). Time-series statistical analysis was carried out using FILM with local autocorrelation correction^49^. The time series model included trial onsets convolved with a double γ HRF function; six motion parameters were added as confound regressors. Indicator functions were added to model out single volumes identified to have excessive motion (>0.9 mm). A separate regressor was used for each high motion volume (deviating more than .9mm from the mean position). For the motor mapping scans, the median number of outlier volumes for an individual scan, across all participants, was 1 volume (range: 0 – 32).

### Low level task-based analysis

We applied a general linear model (GLM) using FEAT to each functional run. Each movement’s task data was modeled against rest resulting in z-statistic maps, where each voxel represents the statistical significance of functional activity relative to rest, with the results thresholded for significance. As previously described, the number of runs performed varied between 1 - 4 across participants, for a given task. For participants with just 1 run of a task, the resulting z-stats were registered to the participant’s structural T1w image. For participants with multiple runs of a single task, estimates from the individual runs were averaged in a voxel-wise manner using a fixed effects model with a cluster forming z-threshold of 2.3 and a family-wise error corrected cluster significance threshold of p < 0.05. Run-averaged z-stats were registed to the participant’s structural T1w scan using FLIRT.

### Cortical surface reconstruction

Structural T1w images were used to reconstruct the pial and white-gray matter surfaces using Freesurfer’s recon-all command (https://surfer.nmr.mgh.harvard.edu/fswiki/recon-all). The grey and white matter segmentations of the reconstructions were then visually inspected and manually edited to correct any large geometric inaccuracies (for more information on the method for performing manual corrections of cortical surface reconstructions, please see https://surfer.nmr.mgh.harvard.edu/fswiki/FsTutorial/PialEdits_freeview). Surface co-registration across hemispheres and participants was done using spherical alignment. Individual surfaces were nonlinearly fitted to a template cortical surface, first in terms of the sulcal depth map, and then in terms of the local curvature, resulting in an overlap of the fundus of the central sulcus across participants^50^.

### Mapping functional activity onto the cortical surface

For surface-based analyses, functional mapping data, already registered to the structural T1w, were then projected onto the cortical surface using the workbench command’s *volume-to-surface-mapping* function which included a ribbon-constrained mapping method. One important consideration is that different tasks were attempted across participants. Therefore, for scans where a single ankle was moved (right or left ankle), we opted to map the activity for only the contralateral hemisphere, as displayed in Figure 2. Alternatively, for scans involving moving both ankles, activity for each hemisphere (where a single hemisphere activity would include both the ipsi- and contra-activations) were mapped onto the cortical surface.

### Regions of interest

Defined regions of interest (ROIs) were generated using one of two approaches: surface-to-surface mapping of the Glasser Human Connectome Project parcellation atlas^51^ and (2) using probabilistic cytoarchitectonic maps^50^. From the Glasser atlas, the following regions were mapped (using *mri_surf2surf*) onto each participant’s cortical surface: M1 (BA4), S1 (BA3b) and medially extended S1 (BA3b, BA5m, BA5mv, BA1). For the univariate ROI analysis, we were interested in activity directly under the SSS. Therefore, we further restricted the left- and right-hemisphere M1 and S1 ROIs to their most medial portions. We also defined a SMA region, which does not have strict anatomical boundaries^52^. Considering the Glasser atlas defines multiple sub-regions on the medial surface, just rostral of M1 [the medial caudal portion of BA6, superior frontal language area (SFL), and supplementary and cingulate eye field (SCEF)], we opted to use the SMA (pre-SMA) region boundaries used in ref^53^, defined using probabilistic cytoarchitectonic maps on the group average cortical surface. These boundaries were then projected onto each participant’s cortical surface. Additionally, as some ROI analyses were performed on the volume, these ROIs were mapped to the volume, using freesurfer’s label-to-volume-mapping method with the ribbon-constrained option.

As a supplementary analysis for P5, we generated the boundaries of the precentral gyrus using the Desiken-Killiany Atlas (https://surfer.nmr.mgh.harvard.edu/fswiki/CorticalParcellation), default to freesurfer. This was displayed in Supp. Figure 1C.

### Univariate analyses

#### Visualizing activation maps

As previously described, for each participant, statistically thresholded ankle activity were mapped onto the cortical surface. In Figure 2A, these activation maps were displayed with a common minimum statistical threshold (Z > 2.3). As a supplementary analysis for 3 participants that underwent additional scans when attempting to move their hands, we displayed the ankle activation maps alongside the hand activation maps (Supp. Figure 1C). Next, we generated a group-level ankle movement activation map, the individual participant activation maps (z-stats) were averaged in a voxel-wise manner. This approach was taken, as opposed to using a mixed-effects model, due to the task differences across participants. The resulting group average z-stat was then mapped onto a standard cortical surface. This activity was minimally thresholded (Z > 2.3) and displayed on a standard pial cortical surface (Figure 2B).

#### Activity within regions of interest

Using the SMA ROI and the medially constrained M1 and S1 ROIs, the average z-stat within each region was extracted for each participant (Figure 2C). Additionally, to estimate the spatial coverage of activity within the regions, the number of significantly activated voxels (Z > 2.3) was computed (Supp. Figure 1B).

### Generating a superior sagittal sinus segmentation

In the investigation, one feature we aimed to extract is spatial information about the SSS. To generate this, we segmented out the neurovasculature in 3D Slicer (https://www.slicer.org/). This was performed using a pipeline that leveraged a Vesselness filtering module, comprehensively described at https://github.com/lassoan/SlicerSegmentationRecipes. The resulting segmentation was then displayed on its own and edited manually to remove secondary vessels. The resulting segmentation was then saved as a .STL file (see Supp. Figure 2).

### Determining the location of the Stentrode BCI

#### Using the post-implant CT image to isolate the Stentrode

All participants underwent post-implant CT scans to determine the position of the Stentrode BCI. CT scans were taken 3-months after implantation. Each participant’s CT image was registered to their structural T1w MRI scan using FSL’s FLIRT (FMRIB’s Linear Image Registration Tool). To ensure the Stentrode image was not atypically morphed, a rigid-body registration was performed (6 degrees of freedom) using the mutual information cost function. Finally, the search range for the rotation parameters were set to [-180°, 180°] along the x, y, and z axes. To extract the location of the Stentrode, the CT-registered image was loaded into 3D Slicer. On a slice-by-slice basis, the Stentrode was manually segmented out for each participant. The resulting segmentation was saved as an .STL file (see Supp. Figure 2).

Next, we displayed the cortical surfaces and segmentations for the SSS and Stentrode in a common coordinate space (see example in Figure 3A). First, we modified the length of the manual Stentrode segmentation. Across participants, the resulting Stentrode segmentations ranged in length from 26.7 – 33.4mm. However, the exact distance of the Stentrode’s most rostral electrode to the most caudal electrode is 25mm. Therefore, we restricted the length of each segmentation from the most rostral point of the segmentation back 25mm. Next, we aimed to generate a simplified approximation of the Stentrode position. We generated a straight line from the most rostral point of the segmentation to the most caudal point of the segmentation. We next wanted to curve this line and position it within the middle of the SSS. We used custom code written in MATLAB, whereby the rostral and caudal line endpoints were positioned in the center of their relative location in the SSS. The midpoint of the line was then similarly adjusted. A polynomial fit was then applied to the line, generating a curved path that approximated the Stentrode’s real-world positioning within the SSS.

To generate the images in Figure 4 of the Stentrode BCI position with the cortical surfaces, a spatially accurate Stentrode model (black) was then aligned to the segmentation such that the most rostral segmentation point coincides with the most rostral point of the Stentrode model. The Stentrode model was then manually positioned to best fit the orientation of the manual segmentation.

### Computing spatial distances

Using the 25mm curved Stentrode line, we defined a point along the line every 0.5mm. Then, we computed the distance from each point to the nearest cortical surface vertex in the left hemisphere and the right hemisphere (50 distance estimations per hemisphere). We displayed a visualization of this approach in Figure 3B. For each hemisphere separately, we computed 5 different distances: 1) the minimum distance of each of these projections to either the left or right hemisphere (i.e., minimum distance to cortex), 2) the average distance to the cortical surface, averaging across the lines for each hemisphere separately (i.e., average distance to cortex), 3) the minimum distance to M1, 4) S1, and 5) SMA. We averaged the distances for each hemisphere to get 1 value per participant. Finally, the width of the SSS was also computed. This was performed by restricting the SSS segmentation to just what was in-line with the Stentrode line and computing multiple distance estimations across the full extent of the SSS segmentation (an example displayed in Supp. Figure 3A).

### Percent overlap of Stentrode and brain regions

To compute a measure of overlap between the Stentrode and brain regions (SMA, M1, S1 etc.), we defined the most rostral point of each region on the medial surface, for each hemisphere separately. A 2-D coronal plane was generated at each point, such that it cut through the Stentrode segmentation. Considering the brain regions are spatially continuous along the medial wall, the boundaries of each region ran from the most rostral point of each region to the most rostral point of the subsequent caudal region (e.g., SMA = rostral of SMA to rostral of M1). The percent overlap values were computed by calculating the distance of Stentrode overlapping each region and dividing these values by the total distance of the Stentrode (25mm). We averaged the percent overlap values across hemispheres to get 1 value per region.

### Cortical thickness

After the Glasser Atlas’^51^ M1 and S1 boundaries were mapped to each participant’s pial cortical surfaces (using a ribbon-constrained manner), the masks were visualized on both the surface and volume to ensure they were spatially accurate (i.e., only included the grey matter). To compute cortical thickness, we implemented the *mri_segstats* command to compute the cortical thickness for each region and hemisphere. We averaged the cortical thickness values across hemispheres to get 1 value per region.

### Computing the volume of the cerebrospinal fluid

To investigate whether the amount of CSF increases with cortical atrophy, we aimed to compute a measure of CSF volume. We usued FreeSurfer’s *mri_segstats* tool to extract total volumes of the CSF, grey matter, white matter and subcortical structures (visualized in Supp. Figure 3B for 1 participant). We then subtracted the SSS segmentation for each participant from the CSF segmentation to yield an adjusted intracranial CSF estimate excluding the primary venous structure.

### Computing a measure of Stentrode motor signal strength

#### Motor signal tests

Throughout both clinical trials, participant engaged in a task to test their ability to generate a click using their Stentrode BCI. The task involved refraining from making a click for 10s (rest period) and attempting the corresponding movement on the screen within 10s (go period), guided by visual cues. There were 10 trials per CCT run and every run finished with an additional 10s rest period (for a breakdown of the number of sessions collected see Supp. Table 3 below). All tests were conducted within the home of each participant.

**Supp. Table 3.** Number of Stentrode motor signal test sessions and body-parts tested across participants. The bolded body-part is the preferred body-part for Stentrode control.

| Subj ID | Body-part tested (number of sessions) | Average number of blocks per session |
| --- | --- | --- |
| P1 | **Left ankle** (3) | 2.33 |
| P2 | **Both ankles** (1), left ankle (4) | 2.20 |
| P3 | **Right ankle** (2) | 1 |
| P4 | **Both ankles** (58), left ankle (2) | 2.25 |
| P5 | **Both ankles** (18) | 2.44 |
| P6 | Both ankles (39), left hand (1), **right hand** (22) | 2.38 |
| P7 | **Both ankles** (10), right ankle (1), both hands (1), right hand (3), right hip (1), mouth (2), left shoulder (1) | 4.73 |
| P8 | **Both ankles** (13), left hand (1) | 4.62 |
| P9 | Both ankles (4), **right hand** (20) | 2.27 |
| P10 | Both ankles (1), **right hand** (19) | 3.26 |

#### Sensitivity index

The sensitivity index was calculated in the following steps. For each participant, channels with high impedance values (>700 kOhms) and exhibiting signs of erratic pertubations via visual inspection were omitted. Data from the remaining channels were bandpass filtered at 100-200 Hz with a 3^rd^ order Butterworth infinite impulse response filter, based on a previous study showing that movement-related modulation in Stentrode recordings is most prominent within this frequency range^14^. Instantaneous power within 100ms non-overlapping windows were derived by calculating the log of the variance per channel, then the values were averaged across the channels. The sensitivity index (*d*′) was calculated between all rest periods and go periods, where:

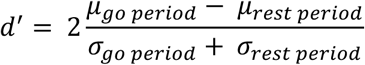

And μ and α denote the mean and the standard deviation of each period. For each participant, their highest sensitivity index value was then used as their Stentrode motor signal strength value.

### Statistical analyses

All statistical analyses were performed using either Python scripts utilizing scipy.stats and statsmodels.stats.multitest or JASP (0.17.2.1). Tests for normality were conducted using a Shapiro–Wilk test. Though, due to the small number of samples (n=10), we opted to primarily use non-parametric statistics in most instances. For the functional neuroimaging data, when comparing whether 1 region had more activity than another region, we used a paired Wilcoxon-signed rank test. For the structural neuroimaging data, when comparing cortical thickness measures between regions (M1, S1) and groups (motor neuron disease, controls), we used a repeated measures ANOVA with region as a within-subject factor and group as the between-subject factor. Post-hoc group comparisons were performed with non-parametric Mann Whitney U Tests where p-values were corrected for 2 comparisons. Additionally, when comparing demographics between the Stentrode participants with motor neuron diseases and the control participants, a Mann Whitney U-test was used to test for differences in age and a chi-squared test was used to test for differences in sex. For the Stentrode motor signal strength data, for each participant, we tested whether values were significantly greater than 0 using a Wilcoxon signed-rank test with 0 set as the test value. For the correlation analyses, we used non-parametric Spearman correlations. Finally, to investigate the relationship between selected predictors and motor signal strength (sensitivity index), we performed a Lasso regression analysis. To standardize the data, we applied a standard scaling procedure to the predictor variables. The Lasso model was fitted with an alpha value of 0.1, which is a regularization parameter that controls the strength of the penalty on the coefficients. A threshold of 0.05 was used for statistical significance.

## Data Availability

The data cannot be made publicly available upon publication because they contain commercially sensitive information. The data that supports the primary findings of this study are available upon reasonable request.

## Acknowledgements

We thank our study participants for their immense generosity and dedication to contributing to this research. We thank Prof. Tamar Makin for sharing the control participant dataset. Finally, we thank Dr. Grace Edwards and Prof. Jessica Taubert for feedback on imaging analyses. For participants P7 and P10, the imaging scans were conducted at the University of Pittsburgh’s Magnetic Resonance Research Center (RRID:SCR_025215).

## Authors contributions

HRS, PY, DJW and JLC conceived the investigation. DJW, DFP, and TJO secured funding. NYH, DFP and DL coordinated clinical management. CHM and FL supported MRI scanning at one of the scan sites. RGN, EL and SM performed the implantation surgeries at one of the sites. HRS led the data analysis with assistance from PY and CH while supervised by JLC. HRS and JLC wrote the paper and all authors (HRS, PY, AF, NC, AS, CH, FL, CHM, KH, SM, NYH, RGN, EL, DFP, DL, TJO, DJW, JLC) critically revised the text and approved the final version.

